# Exploiting collider bias to apply two-sample summary data Mendelian randomization methods to one-sample individual level data

**DOI:** 10.1101/2020.10.20.20216358

**Authors:** Ciarrah Barry, Junxi Liu, Rebecca Richmond, Martin K Rutter, Deborah A Lawlor, Frank Dudbridge, Jack Bowden

## Abstract

Over the last decade the availability of SNP-trait associations from genome-wide association studies data has led to an array of methods for performing Mendelian randomization studies using only summary statistics. A common feature of these methods, besides their intuitive simplicity, is the ability to combine data from several sources, incorporate multiple variants and account for biases due to weak instruments and pleiotropy. With the advent of large and accessible fully-genotyped cohorts such as UK Biobank, there is now increasing interest in understanding how best to apply these well developed summary data methods to individual level data, and to explore the use of more sophisticated causal methods allowing for non-linearity and effect modification.

In this paper we describe a general procedure for optimally applying any two sample summary data method using one sample data. Our procedure first performs a meta-analysis of summary data estimates that are intentionally contaminated by collider bias between the genetic instruments and unmeasured confounders, due to conditioning on the observed exposure. These estimates are then used to correct the standard observational association between an exposure and outcome. Simulations are conducted to demonstrate the method’s performance against naive applications of two sample summary data MR. We apply the approach to the UK Biobank cohort to investigate the causal role of sleep disturbance on HbA1c levels, an important determinant of diabetes.

Our approach can be viewed as a generalization of Dudbridge et al. (*Nat. Comm*. **10**: 1561), who developed a technique to adjust for index event bias when uncovering genetic predictors of disease progression based on case-only data. Our work serves to clarify that in any one sample MR analysis, it can be advantageous to estimate causal relationships by artificially inducing and then correcting for collider bias.

## Background

Mendelian randomisation (MR) is a technique used to test for, and quantify, the causal relationship between a modifiable exposure and health outcome with observational data, by using genetic variants as instrumental variables [1, 2]. MR circumvents the need to measure and adjust for all variables which confound the exposure-outcome association, and is therefore seen as an attractive additional analysis to perform alongside more traditional epidemiological methods [3]. The following Instrumental Variable assumptions are usually invoked in order justify testing for a causal effect of an exposure *X* on a health outcome *Y* using a set of genes, *G*:

- IV1: *G* must be associated with *X*;
- IV2: *G* must be independent of unmeasured confounding between *X* and *Y* ;
- IV3: *G* must be independent of *Y* conditional on *X* and all confounders of the *X*-*Y* relationship.

These assumptions are encoded in the causal diagram in Figure 1. Further linearity and homogeneity assumptions are needed in order to consistently estimate the magnitude of the causal effect. When performing an MR-analysis it is best practice to pre-select SNPs for use as instruments using external data, in order to avoid bias due to the winner’s curse [4]. Subsequently, if the genetic variants are not as strongly associated with the exposure as in the discovery GWAS, assumption IV1 will only be weakly satisfied, which leads to so-called weak instrument bias [5, 6]. This issue is mitigated as the sample size increases as long as the true association is non-zero. When a genetic variant is in fact associated with the outcome through pathways other than the exposure, a phenomenon known as horizontal pleiotropy [7], this is a violation of assumptions IV2 and/or IV3. Horizontal pleiotropy is not necessarily mitigated by an increasing sample size and is also harder to detect. Its presence can therefore render very precise MR estimates hopelessly biased. Pleiotropy-robust MR methods have been a major focus of research in recent years for this reason [8, 9, 10, 11].

**Figure 1:**
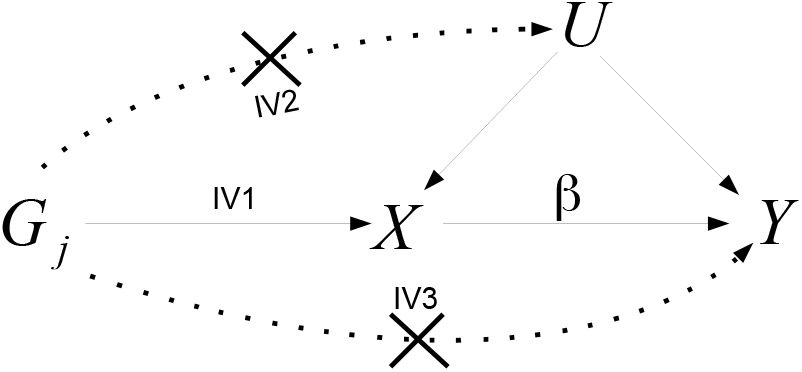
The IV assumptions for a genetic variant *G* are represented by solid lines in the directed acyclic graph (DAG). Dotted lines represent violations of IV assumptions as described in IV2 and IV3. The causal effect of a unit increase of the exposure,*X*, on the outcome,*Y*, is denoted by *β. U* represents unobserved confounders of *X* and *Y*

### One-sample versus Two-sample MR: pros and cons

Obtaining access to a single cohort with measured genotype, exposure and outcome data that is large enough to furnish an MR analysis has been difficult, historically. It has instead been far easier to obtain summary data estimates of gene-exposure and gene-outcome associations from two independent studies, and to perform an analysis within the ‘two-sample summary data MR’ framework (see Figure 2). [12, 13]. This has made it an attractive option for the large scale pursuit of MR, through software platforms such as MR-Base [14]. The relative simplicity of these methods (which resemble a standard meta-analysis of study results) and their ability to furnish graphical summaries for the detection and adjustment of pleiotropy [15] has also acted to increase their popularity. Indeed, the array of pleiotropy robust two sample summary data methods far outstrips those available for one sample individual level data MR analysis [16].

**Figure 2:**
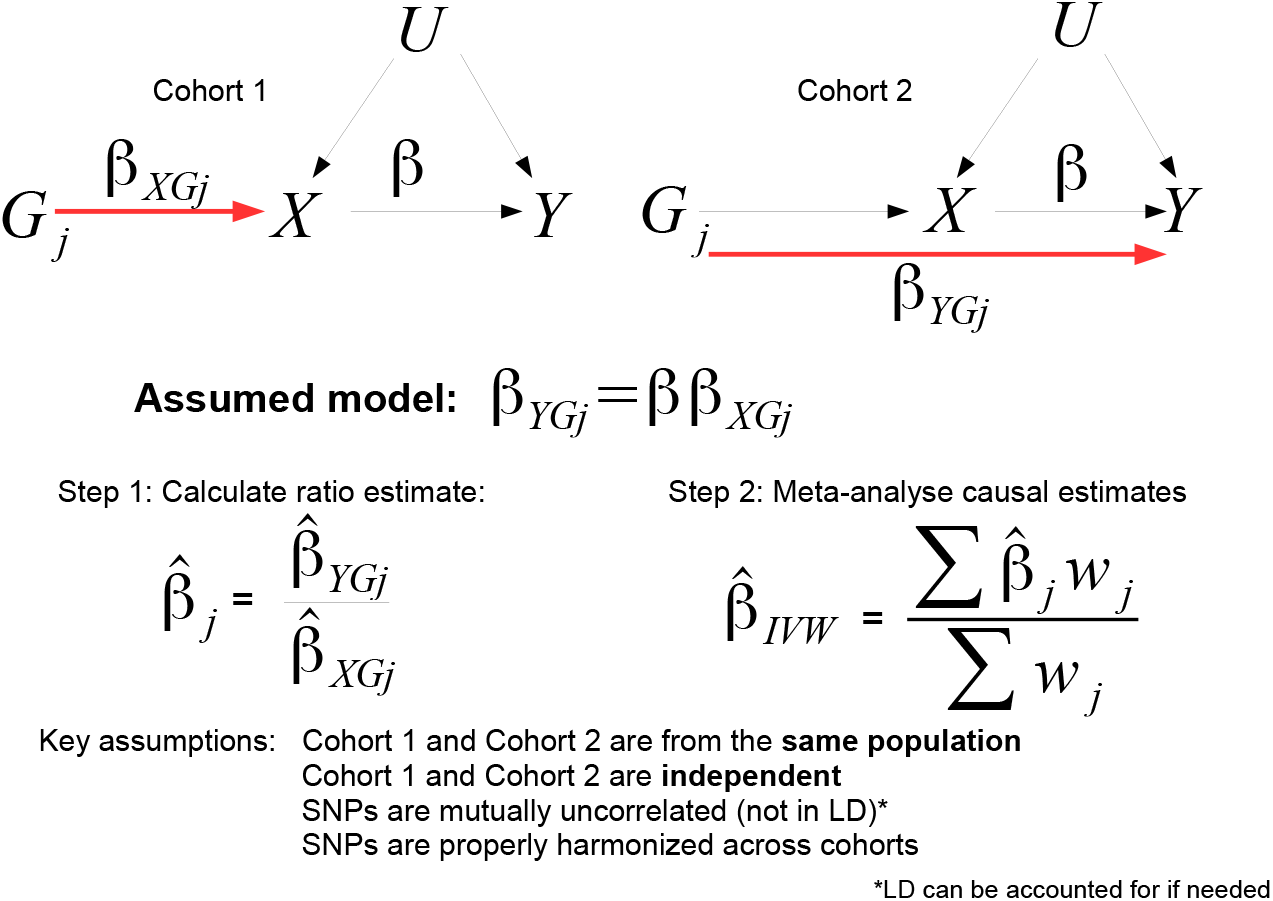
In two sample summary data MR, (*G* − *X*) association estimates, 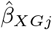, from one cohort are combined with (*G* − *Y*) association estimates, 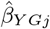 from a separate, non-overlapping cohort, to produce a set of SNP-specific causal estimates, 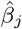. These are combined using inverse variance weighted meta-analysis (*w*_*j*_ being the weight) to obtain an overall estimate 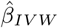 for the true causal effect *β*.

A further advantage of two-sample over one-sample MR is that weak instruments bias causal estimates towards the null (it is often referred to as a ‘dilution’ bias for this reason) which is conservative [17]. Dilution bias arises precisely because uncertainty in the SNP-exposure association estimates obtained from one cohort is independent of the uncertainty in SNP-outcome association estimates from a non-overlapping cohort (Figure 2). This makes the the SNP-exposure association uncertainty akin to ‘classical’ measurement error and enables standard approaches such as Simulation Extrapolation [5, 19] or modified weighting [6, 20] to be used to adjust for its presence. In contrast, weak instruments bias MR estimates obtained from a one sample analysis towards the observational association because uncertainty in the SNP-exposure and SNP-outcome association estimates are correlated. This bias is harder to correct for and is potentially anti-conservative.

There are, however, many disadvantages of using two sample summary data compared to individual level data from a single sample MR, some examples of which are now given: The two-sample approach assumes the two cohorts are perfectly homogeneous [13]. If the distribution of confounders is different between the samples, this can result in severe bias [21]. Alternatively, it may be that the independence assumption is violated due to an unknown number of shared subjects across the two studies [22], which cannot be easily removed [23]. Even when the homogeneity assumption is satisfied, two sample methods can give misleading results if the two sets of associations are not properly harmonized [24]. Often, summary statistics from a GWAS have been adjusted for factors that might bias MR results, and the unadjusted data are not available [25]. It may not be possible to source summary data on the exact population needed for a particular analysis, for example on either men or women only when looking at sex-specific outcomes) [26]. Finally, a richer array of analyses are possible with individual level data. For example, the estimation of non-linear causal effects across the full range of the exposure and the exploration of effect modification via covariates.

It is of course possible to naively apply summary data MR methods to the one-sample context, estimating both the gene-exposure and gene-outcome associations in the same sample, an analysis made increasingly easy by the advent of large open-access cohort studies such as the UK Biobank (UKB) [27]. This avoids problems with synthesising and harmonizing data from separate cohorts, but can result potentially anti-conservative weak instrument bias due to correlated error. A preliminary investigation has found that this naive approach is particularly bad for pleiotropy robust approaches such as MR-Egger regression [28, 29]. So far, there is no consensus on how best to implement summary data approaches in the one sample setting.

In this paper we propose a general method which we term ‘Collider-Correction’ that can reliably apply two-sample summary data MR methods to one-sample data, whilst maintaining the simplicity and appeal of the two-sample approach. Our method builds on the work of Dudbridge et. al. [30], who proposed a method to correct for ‘index event’ (or collider) bias in genetic studies of disease progression, when all subjects included in the analysis have been diagnosed with the disease. In this setting, the analysis is open to contamination from collider bias. Our work serves to clarify that the procedure can be extended to any MR analysis where the aim is to estimate the causal effect, by artificially inducing collider bias in the observational association between *X* and *Y* and then correcting for it. This allows any two sample method to be used in a one sample design, thereby benefiting from the plethora of weak instrument and pleiotropy robust approaches available. We show that this approach is (a) statistically efficient compared to artificially splitting the data in two, and (b) will deliver consistent estimates of the causal effect whenever the assumptions of the underlying two-sample approach are satisfied.

Although our method builds on the work of Dudbridge et al, there are several major differences. Firstly, whilst Dudbridge et al focus on the unbiased estimation of the direct SNP-outcome associations, we treat these as nuisance parameters and focus instead on estimation of the causal effect. Secondly, whilst the underlying method we use is closely related to the approach of Dudbridge et al when the chosen method is MR-Egger regression, our paper shows that the underlying method can actually be applied to any MR method. Thirdly, whereas Dudbridge et al propose a solution to adjust for weak instrument bias within the specific context of an MR-Egger model which relies on the InSIDE assumption [9], we propose the use of a SIMEX procedure that can be applied to any regression model, including robust regression models that do not rely on InSIDE for identification. Of course, some recent two-sample approaches have weak-instrument robust weighting built into them, for example MR-RAPS [20, 31]. In this case, SIMEX adjustment is unnecessary.

A major reason for the emergence of weak instrument and pleiotropy robust two-sample MR methods [6, 20, 31] is the avoidance of winner’s curse [4], by using one discovery GWAS for instrument selection and two additional data sources for the two-sample MR analysis (i.e. a ‘three-sample’ design). Although this removes winner’s curse by design, it generally yields far weaker instruments. In practice, it may be hard to obtain data from three independent, homogenous cohorts to enact the three-sample approach, but a nice property of Collider-Correction is that it can be enacted with two-independent data sources rather than three. In Results, we apply Collider-Correction to 1 sample individual level UK Biobank data to investigate the causal role of sleep disturbance on HbA1c levels, using both overlapping and non-overlapping GWAS data for instrument selection. In the former case winner’s curse is seen to induce a dilution in the MR estimates that is not present in the latter case.

We see three scenarios where our Collider-Correction approach is applicable. Firstly, when interest lies in estimation of the causal effect of an exposure *X* on an outcome, *Y* and only summary data on ‘YadjX’ genetic associations are available (for example, waist/hip ratio adjusted for BMI from the GIANT consortium). The second is when researchers have direct access to individual level patient data. This is likely to become much more common over time as further international biobank studies follow the lead of UKB in opening up data access. Extracting the summary statistics for our approach then enables the efficient implementation of any two-sample method to the data. This is attractive because two-sample methods are currently more numerous than one sample methods, more familiar to researchers and more technically advanced (especially in their ability to adjust for weak instrument bias and pleiotropy). Furthermore, if one additional GWAS is available for instrument selection, Collider-Correction enables winner’s curse, weak instrument bias and pleiotropy to be accounted for using two independent data sets rather than three. The third is when data custodians prefer not to grant direct access to individual level data, but are willing to provide the requisite summary statistics for implementing the Collider-Correction approach, safe in the knowledge that the individual-level data analysis can be performed whilst maintaining data security. Allowing large scale, rapid access to confidential data has obvious benefits to the research community and wider society, as demonstrated through initiatives such as OpenSAFELY [32].

## Methods

To motivate ideas, we assume the following individual level data model for the exposure *X* and continuous outcome *Y* for subject *i*:

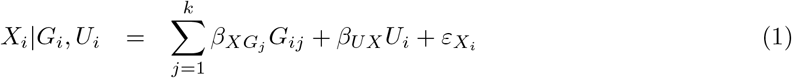

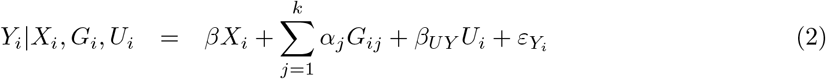

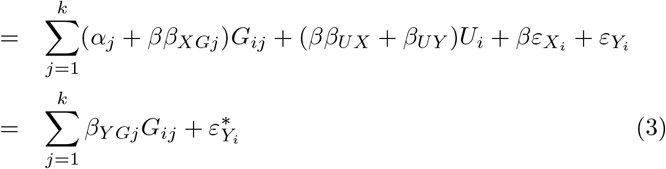

Here, *G*_*i*_ = (*G*_*i*1_, …, *G*_*ik*_) ′ represents a set of *k* variants that predict *X*_*i*_, *β* represents the target estimand, reflecting the causal effect of inducing a 1-unit change in the exposure on the outcome, and *U* represents unmeasured confounding predicting both *X* and *Y*. The variables 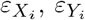 represent independent residual error terms. Since the unmeasured confounder *U* is common to both *X* and *Y*, the total residual errors around *X* |*G, Y* |*X, G* and *Y* |*G* in equations (1) - (3) are correlated. This linear model tacitly assumes that the causal effect is the same for all individuals (that is, regardless of their observed exposure level). This is referred to as ‘Homogeneity’: it is an example of a fourth IV assumption that is needed to ‘point identify’ *β* (assumptions IV1-IV3 are sufficient to test for causality only). We re-write model (2) in ‘reduced form’ as model (3) to clarify that the underlying SNP-outcome association *β*_*Y Gj*_ is equal to *α*_*j*_ + *ββ*_*XGj*_. When the exposure is binary, so that *X*=0, and *X*=1 refer to being unexposed and exposed respectively, we can again identify *β* by assuming Homogeneity. This would mean that the effect of intervening and changing *X* from 1 to 0 is equal and opposite to the effect of intervening and changing *X* from 0 to 1.

The standard approach to estimating *β* with individual level data is Two Stage Least Squares (TSLS). This assumes that all instruments are valid (not pleiotropic), so that *α*_*j*_ = 0 for all *j*. TSLS firstly regresses the exposure on all *k* genotypes simultaneously to derive an estimate for subject *i*’s genetically predicted exposure: 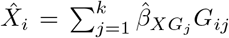, where 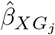 is the estimated association between SNP *j* and *X*. The outcome *Y* is then regressed on 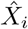 and its regression coefficient is taken as the causal estimate 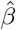. As explained in Figure 2, when the set of *k* SNPs which predict *X* are mutually independent (i.e. not in linkage disequilibrium), the TSLS estimate is asymptotically equivalent to the IVW estimate [33] obtained by:

- Calculating the causal estimate 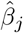 by dividing the SNP-outcome association 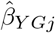 (obtained from a regression of *Y* on *G*_*j*_) by the SNP-exposure estimate 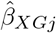 for each SNP and;
- Performing an inverse variance weighted meta-analysis of the *k* individual causal estimates, 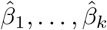.

The inverse variance weights traditionally used make the simplifying assumption that the SNP-exposure association 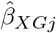 is sufficiently precise that its uncertainty can be ignored. This is referred to as the No Measurement Error (NOME) assumption [6]. This procedure is equivalent to fitting the following weighted regression model

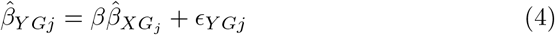

where *ϵ*_*Y Gj*_ is the mean zero residual error with 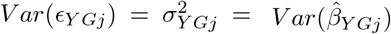 and the intercept is constrained to zero. We will refer to this as the ‘standard’ IVW approach. It is commonly used in two sample summary data MR out of necessity because only summary statistics are available, but not typically in the one sample setting [28].

### Inducing collider bias into SNP-outcome associations

Consider a regression of the outcome *Y* on *G* and *X* together (but not *U*). Under our assumed data generating model:

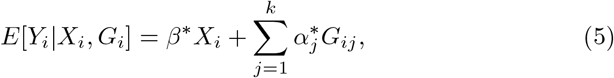

yielding estimated coefficients 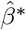 and 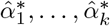. Since *X* is a function of both *G* and *U*, conditioning on *X* induces a correlation between them [34]. This is commonly referred to as ‘collider bias’ [35]. Its presence contaminates the *G*_*j*_-*Y* association estimate with a contribution through *U* so that 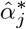 is not a consistent estimate for *α*_*j*_. For the same reason, 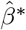 is not a consistent estimate for *β*. It instead reflects the causal effect, plus a contribution from *X* to *Y* via *U*. Such ‘collider biased’ analyses are usually avoided for this reason [35]. However, it is in a special sense advantageous to fit model (5) because under models (1) and (2), 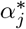, *α*_*j*_, *β*^*^ and *β* are linked through the following linear relation:

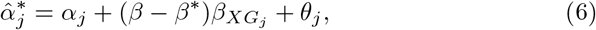

where *θ*_*j*_ is mean zero residual error with 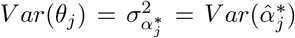 (see the Appendix for a detailed derivation). This suggests the following algorithm for estimating the causal effect:

1. Regress *Y* on *X* and *G* to obtain the collider biased parameter estimates 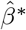 and 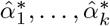.
2. Regress *X* on *G* to obtain estimates 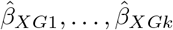, where

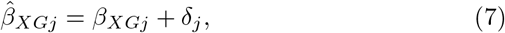

for independent residual error term *δ*_*j*_ with mean zero and variance 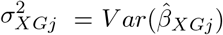;
3. Fit the linear model:

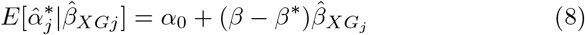

under a user-specified loss function and pleiotropy-identifying assumption in order to obtain an estimate for the Collider-Correction term (*β* − *β*^*^).
4. Adjust the observational estimate to obtain an estimate for the causal effect *β* via:

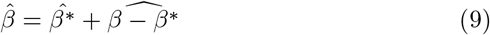

The above procedure, which we call ‘Collider-Correction’ is a modification and generalisation of the Dudbridge approach [30]. In step 3 and 4 we instead focus on estimation of the Collider-Correction term and the causal parameter *β* rather than, as Dudbridge et al do, the pleiotropic effects. Crucially, we clarify that, as long as the model for *Y* given *X* and *G* in step 1 is correctly specified, the correlation between the residual error in model (6) and residual error in the first-stage model (7) will have a mean of zero. To illustrate this we simulated 500 independent sets of data from models (1)-(2), each containing individual level data on 10,000 subjects. We fixed the number of SNPs to *k*=50: each SNP was bi-allelic (taking the values 0,1 or 2), mutually uncorrelated with other SNPs, had a minor allele frequency of 0.3, and collectively explained 1.5% of the variance in the exposure. The correlation between the residual errors in model (1) and (2) was approximately 0.5 to reflect moderate confounding. SNP-exposure and SNP outcome association parameters *β*_*XGj*_ and *α*_*j*_ were generated from dependent distributions, so that their average correlation was approximately 0.45. This is a clear violation of the InSIDE assumption that the sample covariance 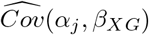 is zero [9] [6, 36]. We then applied Step 1 and 2 of the Collider-Correction algorithm to estimate the 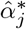 and 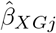 terms. Figure 3 (left) shows, for a single simulated data set, the extent of correlation between the 50 *β*_*XGj*_ and *α*_*j*_. Figure 3 (right) shows across all 500 independent data sets, the sample correlation between the first stage residual 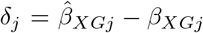 and both:

**Figure 3:**
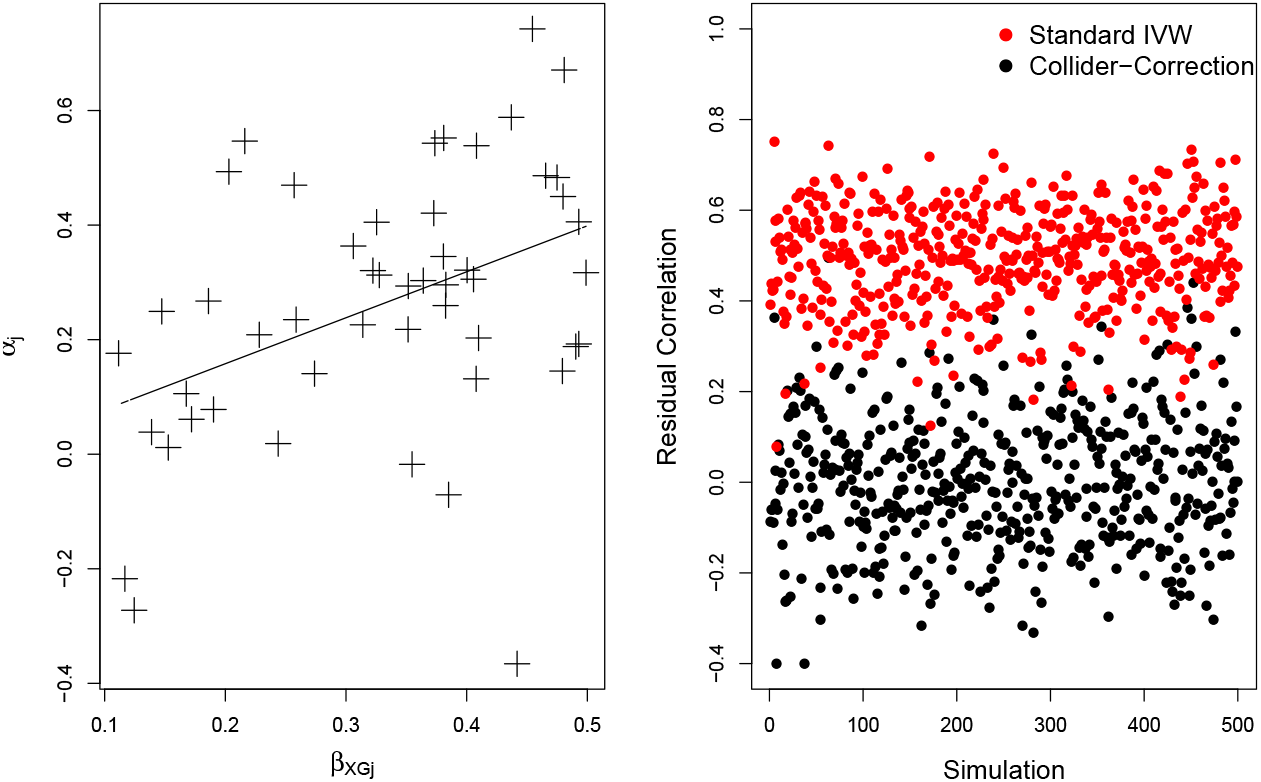
Left: Scatter plot of *β*_*XGj*_ and *α*_*j*_ terms for a single simulated data set. Right Sample correlation between 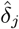 and 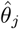 (black) and 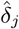 and 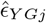 (red).

- The Collider-Correction residual: 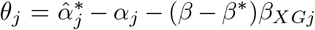 (shown in black);
- The ‘standard’ SNP-outcome residual: 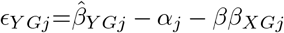 (shown in red).

We see that the mean correlation of the Collider-Correction residual with 1st stage residual is zero whereas the mean correlation of the standard SNP-outcome residual with the 1st stage residual is 0.5.

This residual error independence property is advantageous because it means that step 3 of the Collider-Correction algorithm can be implemented using any pleiotropy robust two-sample summary data MR method, where the estimand of interest is *β* − *β*^*^ rather than the causal effect *β* directly. Crucially, the residual error independence property means that weak instrument bias will induce a dilution in the slope estimate 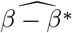 towards zero, because it can be viewed as a consequence of ‘classical’ measurement error. This makes it easy to quantify and correct for using standard methods, as we will subsequently discuss.

In the toy example above we purposefully generated the data so that the InSIDE assumption was violated across the entire set of SNPs to demonstrate that residual error independence does not rely on InSIDE. However, the success of any subsequently applied Collider-Corrected two sample approach in consistently estimating the causal effect *β* (i.e. so that it is asymptotically unbiased) will of course depend on the pleiotropy identifying assumption being met, just as if it were being applied in a standard two-sample setting. Although the Collider-Correction algorithm is generalisable in theory to any MR analysis method, we now describe several canonical implementations, which require that the InSIDE assumption is satisfied across either the entire set of SNPs or a subset of SNPs.

### Implementing Collider-Correction

#### Collider-Corrected IVW implementation

To implement the Collider-Corrected IVW approach we set the parameter *α*_0_ to zero in equation (8) and estimate the slope (*β* − *β*^*^) using weighted least squares via the model:

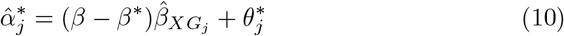

where 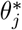 is mean zero residual error with an assumed variance 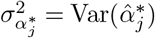. Note that under data-generating model (6) 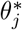 is actually equal to *θ*_*j*_ +*α*_*j*_. Under the assumption that the mean pleiotropic effect is zero and the InSIDE assumption is satisfied, the residual error independence property of Collider-Correction will mean that 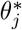 is also independent of uncertainty in 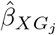 so that (*β* − *β*^*^) can be consistently estimated. The IVW approach then quantifies additional uncertainty in the estimate for (*β* − *β*^*^) due to the presence of pleiotropy, by increasing its variance by a factor *φ* proportional to the variance of the estimated residual 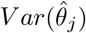 whenever this variance is greater than 1. This is equivalent to fitting a multiplicative random effects model [36].

The IVW estimate uses ‘1st order weights’ that ignore uncertainty in the SNP-exposure association estimate by assuming that its variance 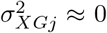. This is referred to as the NO Measurement Error (NOME) assumption [6]. When this is violated the estimate 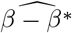 from model (10) will be diluted towards zero by a factor of 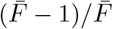, where:

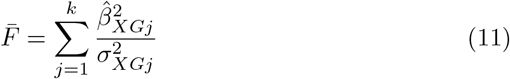

See Section 3.2 in [20] for a more detailed explanation. Note that, whilst the Collider-Correction slope is diluted towards zero in the presence of weak instrument bias, the causal estimate itself is still biased toward the observational association estimate 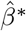, because the causal effect calculated in Step 4 of the Collider-Correction algorithm is the sum of 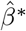 and 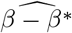. A simple and general method for weak instrument bias adjustment that can be applied directly to the IVW estimate from model (10) is Simulation Extrapolation (SIMEX) [37, 13]. Under SIMEX, a parametric bootstrap is used to generate ‘pseudo’ SNP-exposure associations, each one centred on the observed estimate, but with an increasing amount of uncertainty (i.e. with larger and larger values of 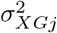). This subsequently induces an increasing dilution in the IVW estimate for (*β* − *β*^*^). A global model is then fitted to the entire set of simulated data in order to extrapolate back to the estimate for (*β* − *β*^*^) that would have been obtained if there were no uncertainty in the SNP-exposure associations (i.e. 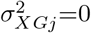, NOME satisfied). SIMEX is attractive because it can be applied to any regression model (and hence many MR methods), and reliable software is available in standard software packages, such as R and Stata.

#### Connecting IVW to LIML and MR-RAPS

An alternative to SIMEX in the special case of the IVW approach is to find the values of (*β* − *β*^*^) and 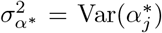 that minimises the weighted sum of squared residuals in the extended model (12):

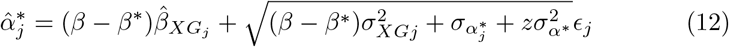

When *z*= 0 in (12), the pleiotropy variance 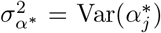 is fixed to zero and the above procedure is equivalent to performing Limited Information Maximum Likelihood (LIML) with summary data (see Section 3.1 in [20]). Furthermore, the weighted sum of squared residuals from (12) follows a 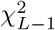 distribution when the assumption that 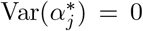 is satisfied, thus providing a simple weak instrument bias robust test for the presence of pleiotropy. This is referred to as the ‘exact’ Q statistic [6] which is similar to the simulation-based MR-PRESSO test for ‘global’ pleiotropy [38, 39].

Unfortunately, when pleiotropy *is* present so that 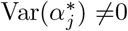, then the LIML estimate will be biased [6]. In order to account for both weak instrument bias and non-zero pleiotropy, *z* can be set to 1 so that the squared residual minimisation is over both (*β* − *β*^*^) and 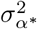. This is equivalent to applying ‘MR-RAPS’ [20] when applied to the Collider-Correction summary statistics. MR-RAPS actually uses an approximation to the least-squares method because the maximum likelihood estimates are inherently unstable, this entails the use of a score function to proxy for the likelihood and a penalization term to dampen the effect of large residuals.

### Collider-Corrected MR-Egger implementation

In order to account for pleiotropy with a non-zero mean but under the InSIDE assumption, we could instead allow the intercept *α*_0_ and slope (*β* − *β*^*^) to be freely estimated via weighted least squares by fitting a Collider-Correction MR-Egger model [9]

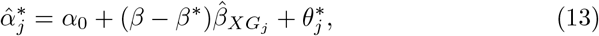

where 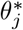 is mean zero residual error with an assumed variance 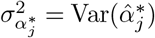. Note that under data-generating model (6) 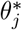 is actually equal to *θ*_*j*_ + *α*_*j*_ − *α*_0_. Using the same argument as for the IVW model, when InSIDE is satisfied this will consistently estimate the Collider-Correction slope (adjusted for *α*_0_) and from there, the causal effect. Additional uncertainty due to pleiotropy can again be handled using a multiplicative random effects model [36]. To assess the vulnerability of the MR-Egger regression estimates to weak instrument bias due to violation of the NOME assumption, we use the 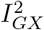 statistic [5]:

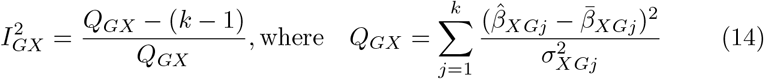

The expected dilution in the Collider-Correction 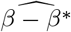 due to weak instruments is equal to 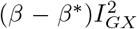. This can easily be adjusted for by applying SIMEX to model (13), just as for the IVW approach.

### Collider-Corrected robust regression

IVW MR-Egger and MR-RAPS rely on the InSIDE assumption to consistently estimate the causal effect. This may be violated in practice, hence the rationale for the development of alternative, robust methods such as the Weighted Median [10]. In the two-sample summary data context it can consistently estimate the causal effect if the majority of the ‘weight’ in the MR analysis stems from genetic variants that are not pleiotropic. That is, the existence of a SNP subset *S* is assumed for which 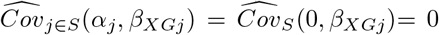, but InSIDE is allowed to be violated for SNPs not in subset *S*. The downside of weighted median approach is that it is not directly equivalent to a regression model, which in turn means that we can not benefit from a procedure like SIMEX to perform a weak instrument bias adjustment. However, there is a close connection between the median and minimisation using a Least Absolute Deviation (LAD), or L1-norm. We therefore propose the use of weighted LAD regression [40] instead of least squares, at Step 3 of the Collider-Correction algorithm, with *α*_0_ set to zero. This is close in spirit to the Weighted Median, and is amenable to SIMEX-adjustment too. The exact ‘breakdown point’ of LAD regression (or the proportion of pleiotropic SNPs above which LAD regression will not deliver a consistent estimate) depends on the data generating model, but is bounded between 1/k (k being the number of SNPs) and 1/2.

### Simulation studies

In order to confirm our theoretical results and assess the performance of the Collider-Correction algorithm, data sets of between 5000 and 50,000 individuals were generated under models (1) and (2) as described previously. Across all simulations:

- The causal effect of inducing a one-unit change in the exposure on the outcome, *β*, was set to 0.5 for all individuals;
- The correlation between the residual errors in model (1) and (2) was set to approximately 0.9 to reflect strong confounding;
- The observational estimate for 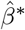 and the true Collider-Correction term *β* − *β*^*^ were approximately 1.12 and -0.62 respectively.

To showcase the ability of IVW-based approaches, MR-Egger regression and LAD regression, pleiotropy parameters and SNP-exposure associations were generated under three distinct models:

- For IVW simulations, pleiotropic effect parameters *α*_1_, …, *α*_50_ were generated with a zero mean independently of the SNP-exposure associations *β*_*XG*1_, …, *β*_*XG*50_ (InSIDE satisfied);
- For MR-Egger simulations, pleiotropic effect parameters *α*_1_, …, *α*_50_ were generated with a non-zero mean independently of the SNP-exposure associations *β*_*XG*1_, …, *β*_*XG*50_ (InSIDE satisfied);
- For LAD regression simulations, pleiotropic effect parameters *α*_1_, …, *α*_15_ were generated with a non-zero mean dependent on the SNP-exposure associations *β*_*XG*1_, …, *β*_*XG*15_ (with an average correlation of 0.5) whilst *α*_16_, …, *α*_50_ were set to 0. InSIDE was therefore strongly violated across SNPs 1:15, satisfied across SNPs 16:50 and violated across all SNPs, respectively.

#### IVW simulation results

Figure 4 (top-left) shows, for a range of sample sizes the average value across 1000 independent data sets of : (a) The standard IVW estimate (black line); (b) the SIMEX adjusted standard IVW estimate (blue line); (c) the Collider-Corrected IVW estimate (red line); (d) the Collider-Corrected IVW estimate with SIMEX correction (green line); (e) the TSLS estimate (orange line) and (f) the Collider-Corrected MR-RAPS estimate (implemented using the ‘Tukey’ penalization option). We see that methods (a), (c) and (e) give approximately the same answer, and are therefore hard to individually distinguish in the figure. The approximate equivalence of the TSLS and IVW approaches with uncorrelated SNPs is well known, but it is also reassuring that our two step approach is also equivalent. We also see that applying a direct SIMEX correction to method (a) (i.e. method (b)) dramatically increases the bias of the causal estimate beyond even that of the observational estimate for small sample sizes. This bias is slow to diminish as the sample size grows. This poor performance is because uncertainty in the SNP-exposure association estimates can **not** be viewed as classical measurement error within a standard IVW model. Conversely, we see that applying a SIMEX correction to the Collider-Corrected IVW estimate (c) (i.e method (d)) yields a steadily decreasing bias which is essentially zero when the mean *F* statistic across the instruments is larger than 5. The Collider-Corrected MR-RAPS estimate performs very well too, and is essentially unbiased for mean *F* statistics greater than 3.5.

**Figure 4:**
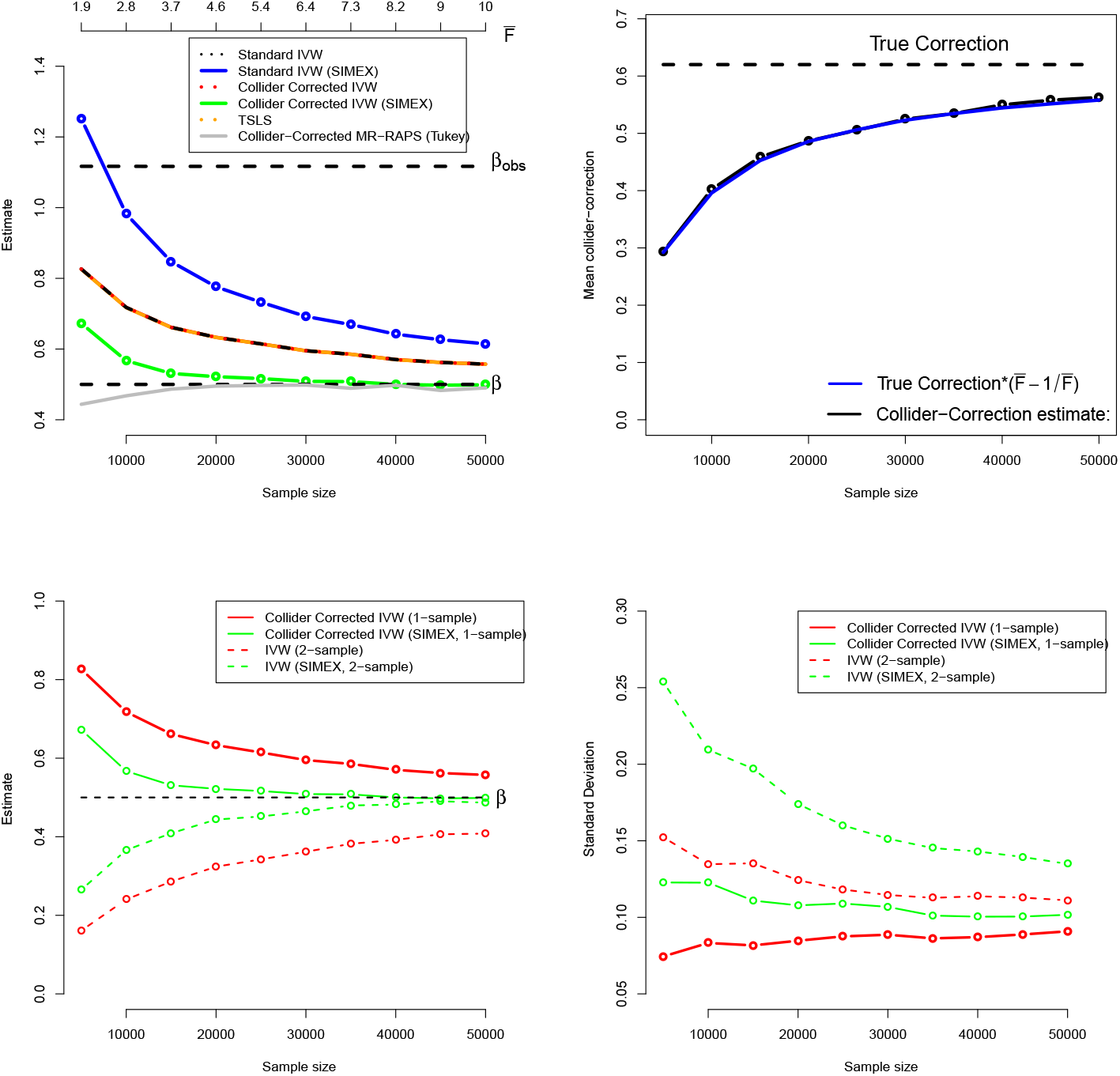
Top: Performance of IVW implementations (including the Collider-Correction algorithm) using one-sample data. Bottom: comparison of the one sample Collider-Correction versus two-sample IVW approaches in terms of bias (bottom-left) and efficiency (bottom-right).

Figure 4 (top-right) gives further intuition on why the correction process works. The black line shows the estimated Collider-Correction 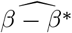 as a function of the given sample size. The blue line shows the true Collider-Correction multiplied by the expected dilution factor 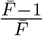, which varies as a function of the sample size. The fact that the two lines are in good agreement indicates that the dilution in 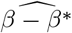 can be perfectly predicted by the F-statistic formula and underlines why SIMEX can be used to correct for it.

Figure 4 (bottom-left) shows the performance of the IVW estimate implemented using the (one sample) Collider-Correction algorithm, versus that obtained from artificially splitting the data in two, and applying the ‘standard’ IVW approach. That is, calculating SNP-exposure associations in one half, SNP-outcome associations in the other half and combining in the usual manner. This ensures that the residual error independence property is satisfied, as it is for the one sample Collider-Correction approach. Results for each method are shown with and without SIMEX correction. We see that the absolute bias of the Collider-Correction implementations is less than that of the two-sample implementation. However, the two estimation strategies differ more substantially in terms of precision, as shown in Figure 4 (bottom-right). Collider-correction of one sample data is shown to be far more efficient than sampling splitting.

Figure 7 and Figure 8 (top-left) in the Appendix show the corresponding standard deviation and mean-squared error (a measure of accuracy that equals an estimate’s variance plus the squared bias) for all IVW-based methods across the same set of simulations. They show that whilst the MR-RAPS estimate is less biased for small sample sizes than the Collider-Corrected IVW method with SIMEX adjustment, it is more variable and less accurate.

#### MR-Egger simulation results

Figure 5 (top-left) shows for a range of sample sizes the average value across 1000 independent data sets of : (a) The standard (one-sample) MR-Egger estimate (black line); (b) the SIMEX adjusted standard MR-Egger estimate (blue line); (c) the Collider-Corrected MR-Egger estimate (red line) and (d) the Collider-Corrected MR-Egger estimate with SIMEX correction (green line). As the sample size increases the 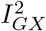 statistic increases from 0.1 to 0.5. This signals that the 50 SNPs get collectively stronger as a set of instruments within MR-Egger as the sample size increases, but even at the largest sample size we expect a dilution of 50% in the MR-Egger slope. Again, we see that standard and Collider-Corrected MR-Egger methods give the same results, but the two approaches differ greatly under SIMEX correction, with the SIMEX adjusted Collider-Corrected estimate being least biased. In Figure 5 (top-right) we show how dilution in the Collider-Corrected slope estimate 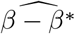 for MR-Egger can be accurately quantified using the 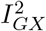 statistic, just as the *F* -statistic predicts the dilution for IVW. This explains why SIMEX adjustment works.

**Figure 5:**
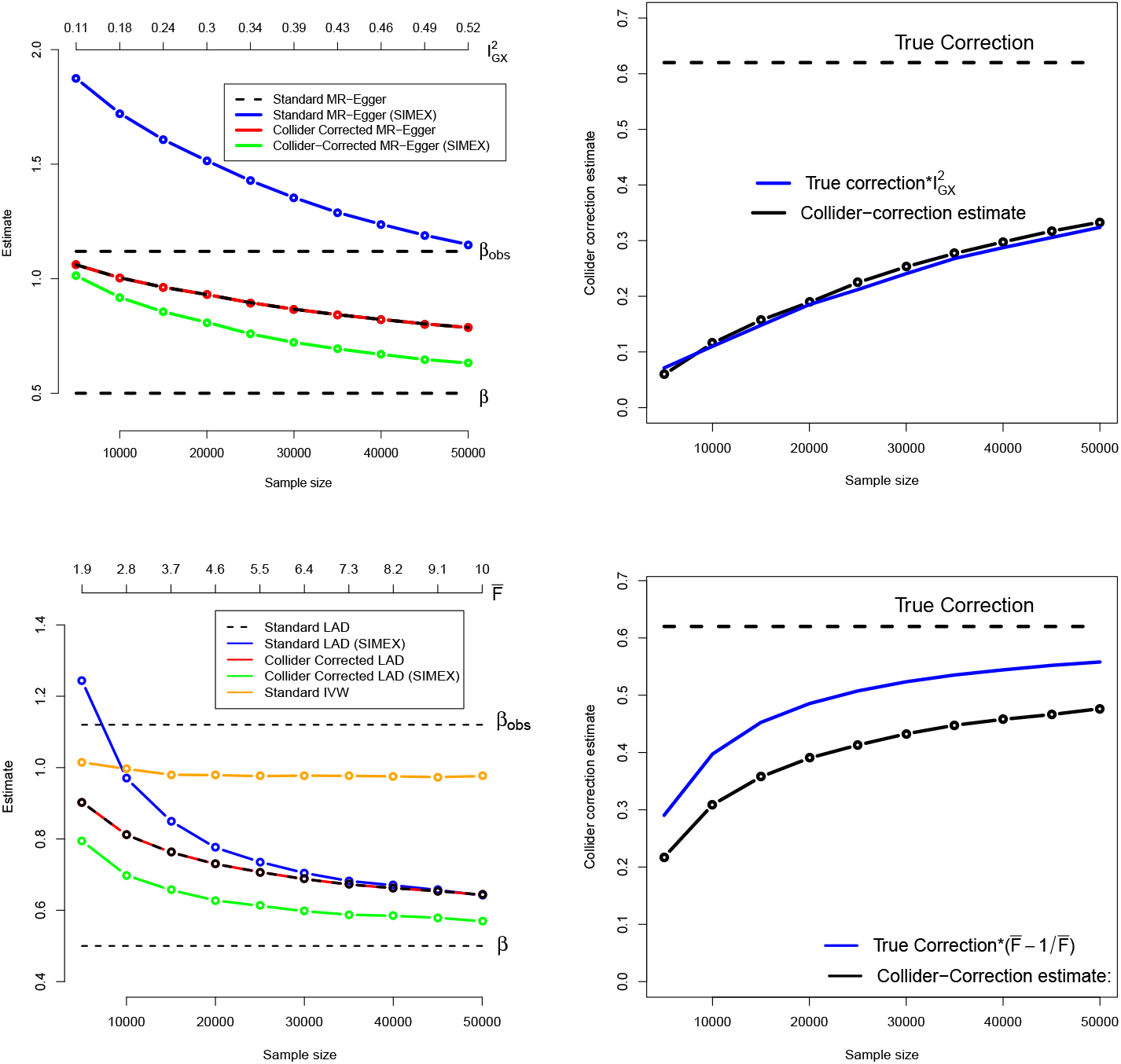
Top: Performance of the MR-Egger implementation of the Collider-Correction algorithm under a directional pleiotropy scenario. Bottom: Performance of the LAD-regression implementation

Figure 7 and Figure 8 (top-right) in the Appendix show the corresponding standard deviation and mean squared error for all MR-Egger-based methods across the same set of simulations. Standard and Collider-Corrected MR-Egger are seen to have the joint smallest variance, but Collider-Corrected MR-Egger with SIMEX adjustment has the smallest mean-squared error because it is far less biased.

#### LAD-regression simulation results

Figure 5 (bottom-left) shows for a range of sample sizes the average value across 1000 independent data sets of : (a) The standard (one-sample) LAD-regression estimate (black line); (b) the SIMEX adjusted standard LAD regression estimate (blue line); (c) the Collider-Corrected LAD regression estimate (red line) and (d) the Collider-Corrected LAD regression estimate with SIMEX correction (green line). For comparison we also show (e) the standard IVW estimate: its bias does not approach zero as the sample size increases because of the presence of non-zero mean pleiotropy violating InSIDE, which is the very motivation for LAD regression. As in the previous simulations, standard and Collider-Corrected LAD regression give identical point estimates on average, but when SIMEX adjustment is applied the two estimates diverge substantially. Collider-Corrected LAD regression with SIMEX adjustment results in the least biased estimates of all.

Figure 5 (bottom-right) plots the mean dilution in the Collider-Corrected LAD regression estimate, versus that predicted by the IVW dilution factor 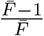. The fact that the observed dilution is below the expected IVW dilution illustrates that LAD regression is more vulnerable to weak instrument bias, because it is a less efficient but more robust technique. This emphasises the importance of being able to address its weak instrument bias.

Figure 7 and Figure 8 (bottom) in the Appendix show the corresponding standard deviation and mean-squared error for all LAD regression-based methods across the same set of simulations. The same pattern of higher variance but lower mean-squared error is seen for the SIMEX adjusted Collider-Corrected LAD regression approach as in the MR-Egger case.

The MR-RAPS approach can, in theory, consistently estimate the causal effect when a small proportion of SNPs are pleiotropic and violate the InSIDE assumption, as long as their contribution is strongly penalized by its robust loss function. In order to test this we also calculated the MR-RAPS estimate when applied to the simulated data for LAD regression. MR-RAPS was seen to work well for a proportion of simulated data sets, but its estimates were unstable: in many cases they were an order of magnitude larger than the true value of 0.5. To illustrate this, Figure 9 in the Appendix shows the distribution of its estimates at the largest sample size of 50,000 subjects, where it was most stable. Even in this case substantial instability is observed.

## Results: Assessing the causal role of Insomnia on HbA1c

Observationally, sub-optimal sleep (i.e., low sleep quantity and quality) has been found to be associated with hyperglycaemia [41, 42, 43] and increased diabetes risk [44]. Insomnia, defined as difficulty initiating or maintaining sleep, is one of the most important indices of sleep quality [45]. It has been associated with type 2 diabetes in observational studies [45] and in a previous Mendelian randomization study [46]. However, it is unclear whether associations with insomnia are mediated through HbA1c in the general population, whose glucose levels may not meet the threshold criteria for a formal diabetes diagnosis. As such, we focus on a potentially causal role of insomnia on HbA1c, a well-established clinical assessment of long-term glycaemic regulation that is central to the diagnosis of diabetes [47]. To address this question we use individual level data on approximately 320,000 individuals in UK Biobank to furnish a one sample Mendelian randomization study.

Two hundred and forty-eight independent genetic variants at 202 loci were associated with self-reported insomnia at or below the standard genome-wide significance threshold (p-value<5 × 10^−8^) in a recent GWAS of over 1.33 million UK Biobank and 23andMe individuals reported by Jansen [46] which collectively explained 2.6% of the total trait variance. SNP-exposure associations were measured on the log-odds scale using logistic regression. Among this set of variants, 240 SNPs were in principle available for use as instruments in UK Biobank. In this cohort, participants were asked: “Do you have trouble falling asleep at night or do you wake up in the middle of the night?” with responses “Never/rarely”, “Sometimes”, “Usually”, or “Prefer not to answer”. Those who responded “Prefer not to answer” were set to missing. To reflect the Jansen analysis, the remaining entries were treated as a binary variable for insomnia symptoms, with “Never/rarely”, “Sometimes”, and “Usually” coded as 0, 0, and 1, respectively and a logistic regression performed. HbA1c measurements were obtained from a panel of biomarkers assayed from blood samples collected at baseline from UK Biobank participants. HbA1c (mmol/mol) was measured in red blood cells by HPLC analysis using Bio-Rad VARIANT II Turbo and log-transformed.

### Instrument selection and winner’s curse

The mean *F* statistic for the 240 genetic instruments in the original GWAS was 41, but in order to avoid winner’s curse we did not want to incorporate these estimates directly into our MR analysis. In UK Biobank the same SNPs had an 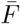 of approximately 8.3 and an 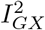 statistic of approximately 40%, meaning that the MR analysis was susceptible to bias due to both weak instrument and pleiotropy. This motivates the use of our Collider-Correction method for causal estimation. However, the original Jansen GWAS combined data from the UK Biobank (n=386,533) and 23andMe (n=944,477) using METAL [48]. As such, there was an approximate 23% overlap between data used for SNP discovery and for estimation in our MR model [4]. To additionally assess the impact of winner’s curse for this reason we performed our subsequent analysis using (a) all 240 SNPs and (b) a subset of 112 SNPs that were only genome-wide significant using only the 23andMe portion of the Jansen data. Analysis (b) is completely protected from winner’s curse whereas (a) is not. The downside of analysis (b) is that, with an 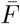 of 6.8, it is even more susceptible to weak-instrument bias.

### Methods used

We applied the TSLS, IVW, MR-Egger, LAD regression and MR-RAPS approaches to the data. The IVW, MR-Egger and LAD regression approaches were implemented in three ways (1) The ‘Standard’ 1-sample approach (i.e. using all the data to estimate SNP-exposure and SNP-outcome associations); (2) the Collider-Correction algorithm and (3) Collider-Correction with SIMEX adjustment. Note that MR-RAPS incorporates an internal weak instrument bias adjustment and there is no need to additionally apply a SIMEX algorithm to it. Along with MR-RAPS, we refer to approach (3) as the ‘gold-standard’ methods.

### Causal estimates

SNP exposure associations 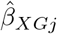 were obtained from a logistic regression of insomnia on the set of SNPs as well age at recruitment, sex, assessment centre, 10 genetic principal components, and genotyping chip. Estimates for collider biased SNP outcome associations 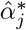 were obtained from a multivariable regression of HbA1c on observed insomnia severity, all genetic variants and the same additional covariates. This second regression additionally yielded an estimate for the collider biased observational association between insomnia severity and HbA1c of 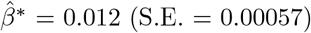 (S.E. = 0.00057).

Figure 6 (top) plots the collider biased SNP-outcome associations versus the SNP-exposure associations for analysis (a). Overlaid on the plot are the weak-instrument and pleiotropy adjusted Collider-Correction slopes 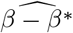 estimated by the four gold standard methods. The *Q* statistic is 809 (df = 239) providing overwhelming evidence of heterogeneity due to pleiotropy. The 13 SNPs circled in black contribute a component to this global statistic with a bonferroni corrected p-value below (5/240)% and could therefore be classed as outliers. Adjusted causal effect estimates can be found in Table 1.

**Figure 6:**
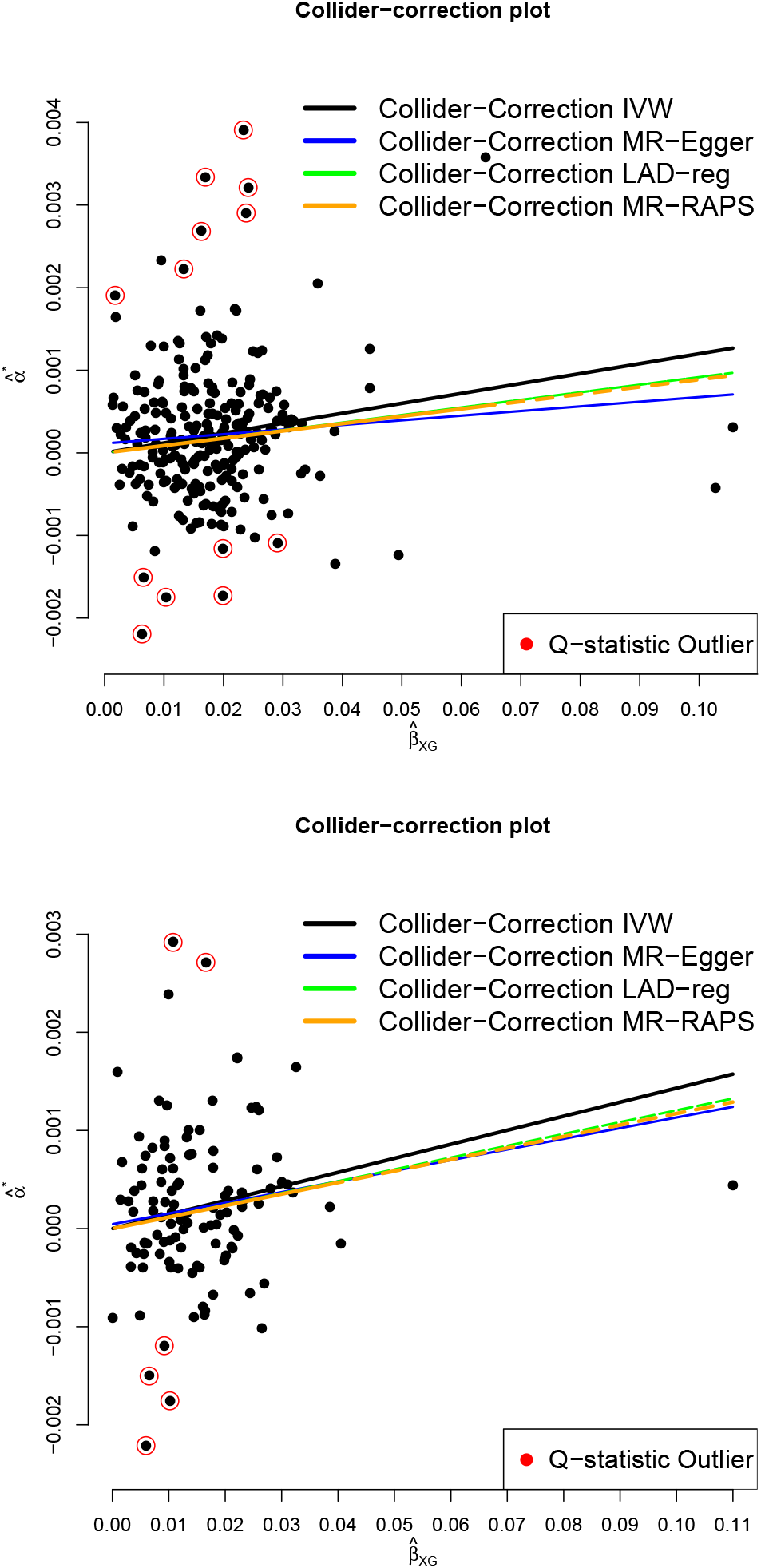
Collider biased SNP outcome associations, 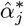, versus SNP-exposure associations, 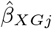 for: 240 SNPs that were genome-wide significant using 23andMe and UKB data (top); the 112 SNPs that were genome-wide significant using 23andMe data only (bottom)

**Figure 7:**
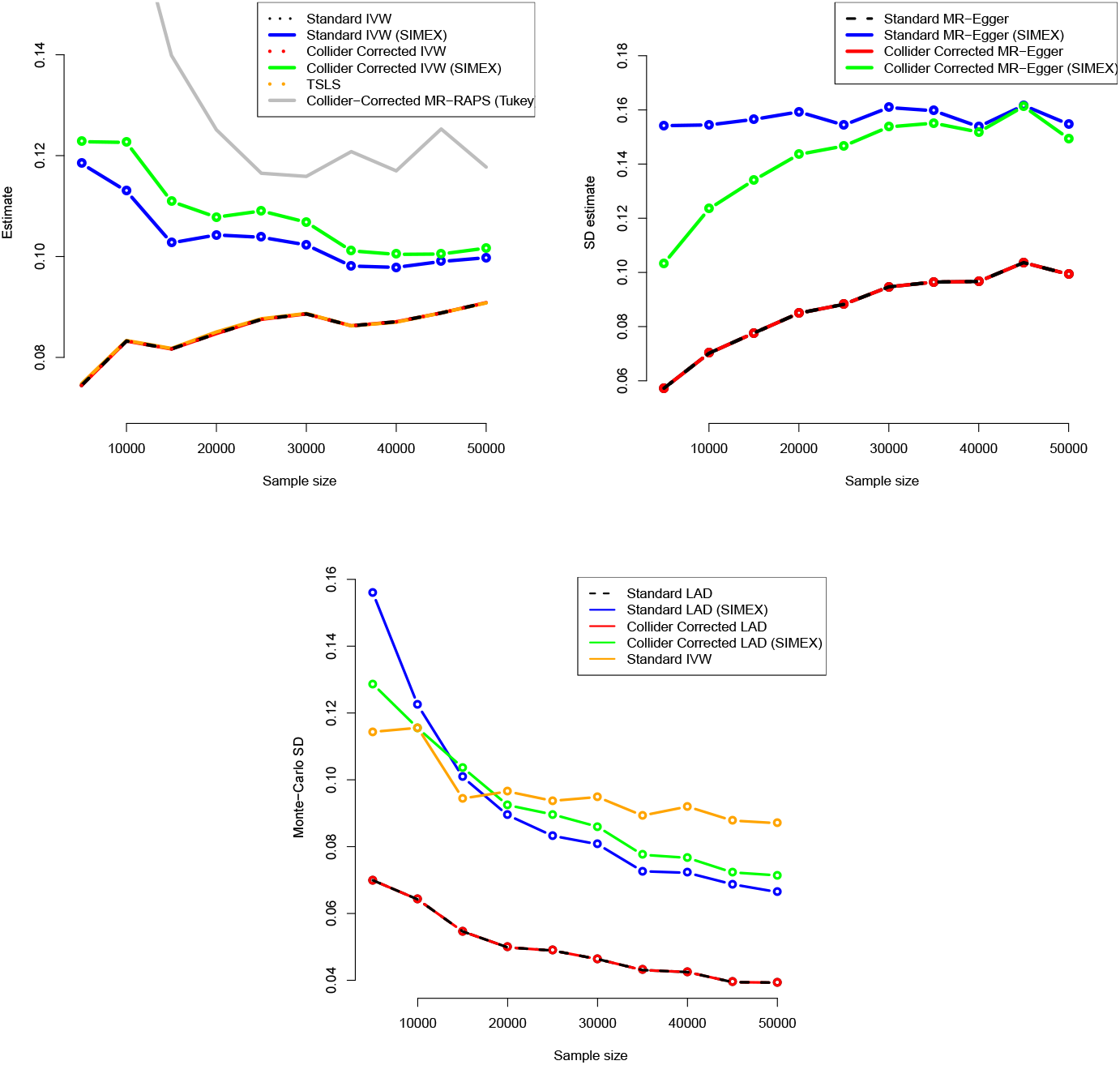
Monte-Carlo standard deviations for all IVW (top-left), MR-Egger (top-right) and LAD regression (bottom) estimators

**Figure 8:**
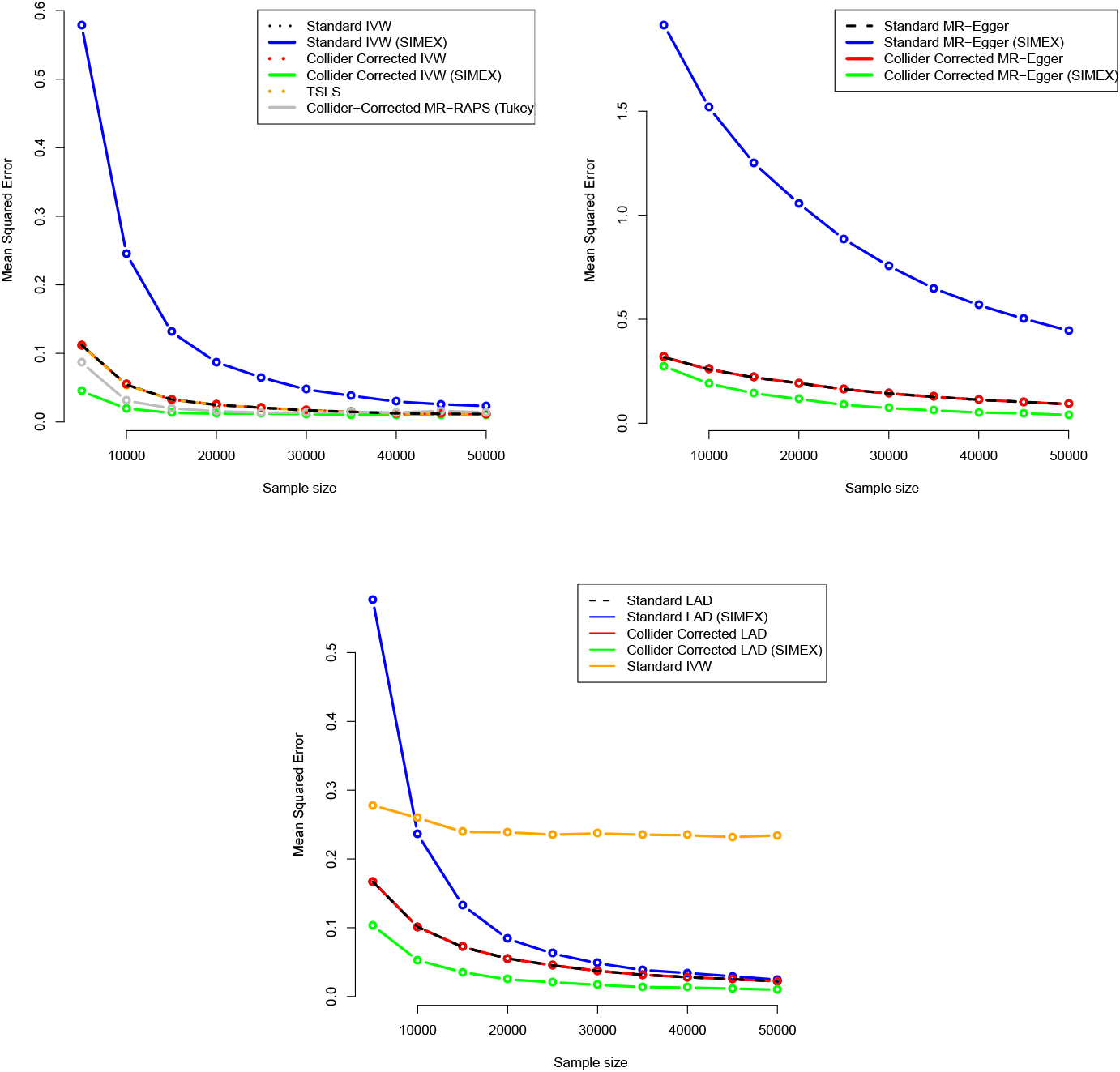
Mean Squared Error for all IVW (top-left), MR-Egger (top-right) and LAD regression (bottom) estimators

**Figure 9:**
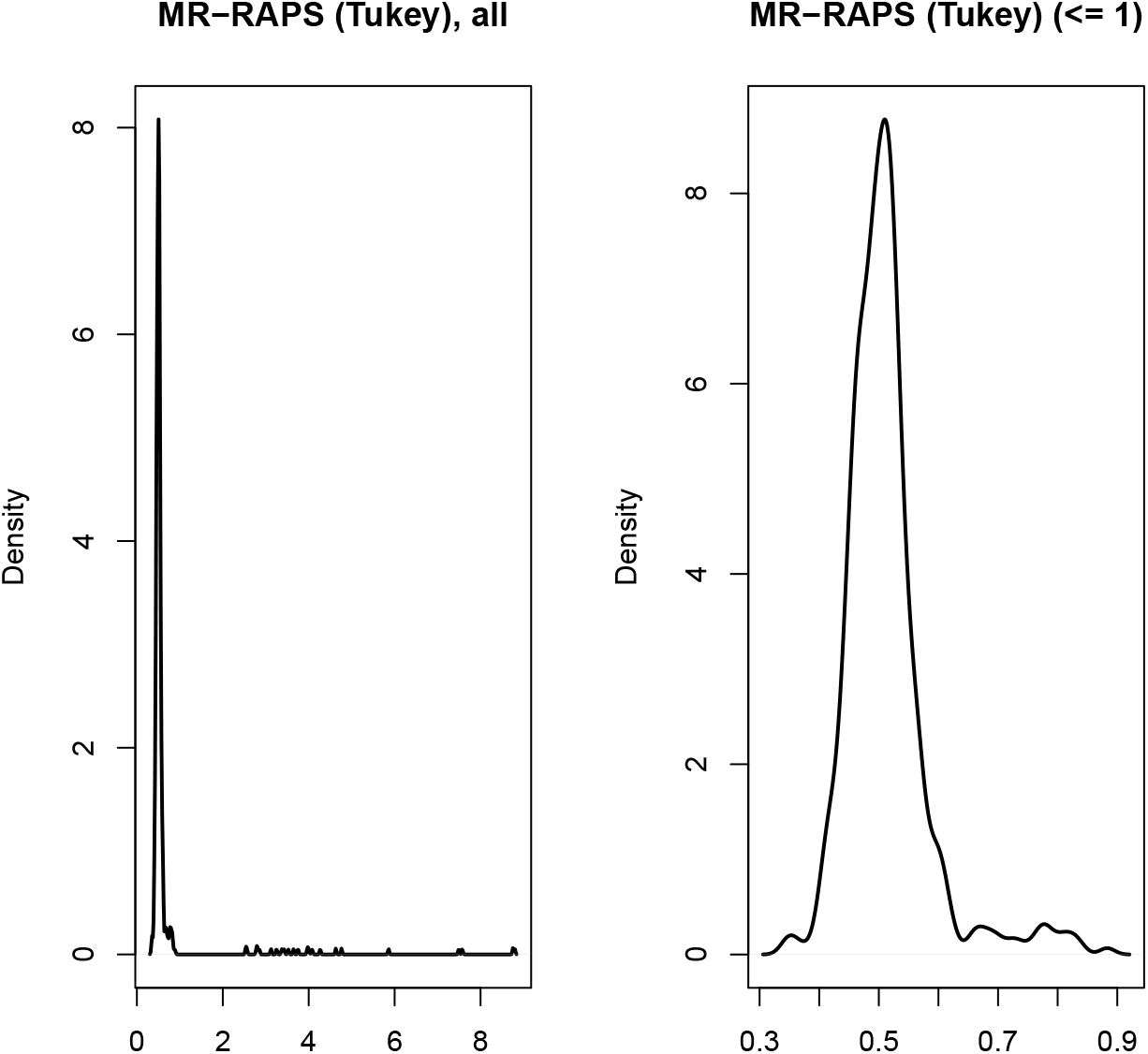
Distribution of MR-RAPS estimates at sample size = 50,000 when the data were generated under the LAD regression model. Left: all estimates, Right: estimates less than 1.

**Table 1:**
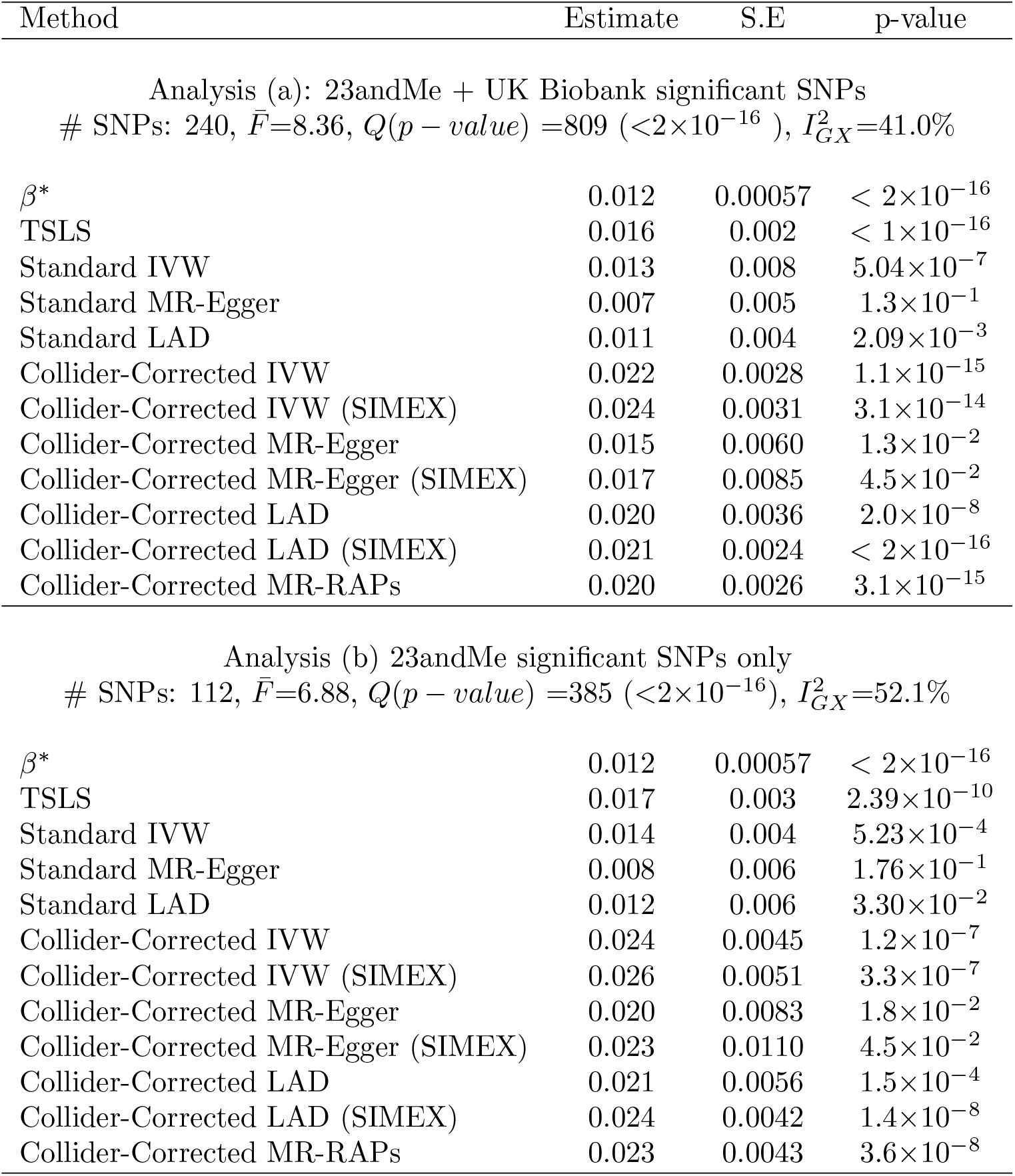
Point estimates, standard errors and p-values for the: TSLS, IVW, MR-Egger, LAD-regression and MR-RAPS methods. Estimates reflect the average causal effect of a unit increase in the log-odds of insomnia on HbA1c levels across the population. ‘Standard’ = standard 1-sample analysis. Top rows: Analysis (a) - All 240 SNPs from Jansen et al used. Bottom rows: Analysis (b) - only genome wide significant SNPs from 23andMe data (ignoring UK Biobank) used.

| Method | Estimate | S.E | p-value |
| --- | --- | --- | --- |
| Analysis (a): 23andMe + UK Biobank significant SNPs<br># SNPs: 240, $\bar{F}$=8.36, $Q(p-value) = 809$ ($< 2 \times 10^{-16}$), $I_{GX}^2 = 41.0\%$ | | | |
| $\beta^*$ | 0.012 | 0.00057 | $< 2 \times 10^{-16}$ |
| TSLs | 0.016 | 0.002 | $< 1 \times 10^{-16}$ |
| Standard IVW | 0.013 | 0.008 | $5.04 \times 10^{-7}$ |
| Standard MR-Egger | 0.007 | 0.005 | $1.3 \times 10^{-1}$ |
| Standard LAD | 0.011 | 0.004 | $2.09 \times 10^{-3}$ |
| Collider-Corrected IVW | 0.022 | 0.0028 | $1.1 \times 10^{-15}$ |
| Collider-Corrected IVW (SIMEX) | 0.024 | 0.0031 | $3.1 \times 10^{-14}$ |
| Collider-Corrected MR-Egger | 0.015 | 0.0060 | $1.3 \times 10^{-2}$ |
| Collider-Corrected MR-Egger (SIMEX) | 0.017 | 0.0085 | $4.5 \times 10^{-2}$ |
| Collider-Corrected LAD | 0.020 | 0.0036 | $2.0 \times 10^{-8}$ |
| Collider-Corrected LAD (SIMEX) | 0.021 | 0.0024 | $< 2 \times 10^{-16}$ |
| Collider-Corrected MR-RAPs | 0.020 | 0.0026 | $3.1 \times 10^{-15}$ |
| Analysis (b) 23andMe significant SNPs only<br># SNPs: 112, $\bar{F}$=6.88, $Q(p-value) = 385$ ($< 2 \times 10^{-16}$), $I_{GX}^2 = 52.1\%$ | | | |
| $\beta^*$ | 0.012 | 0.00057 | $< 2 \times 10^{-16}$ |
| TSLs | 0.017 | 0.003 | $2.39 \times 10^{-10}$ |
| Standard IVW | 0.014 | 0.004 | $5.23 \times 10^{-4}$ |
| Standard MR-Egger | 0.008 | 0.006 | $1.76 \times 10^{-1}$ |
| Standard LAD | 0.012 | 0.006 | $3.30 \times 10^{-2}$ |
| Collider-Corrected IVW | 0.024 | 0.0045 | $1.2 \times 10^{-7}$ |
| Collider-Corrected IVW (SIMEX) | 0.026 | 0.0051 | $3.3 \times 10^{-7}$ |
| Collider-Corrected MR-Egger | 0.020 | 0.0083 | $1.8 \times 10^{-2}$ |
| Collider-Corrected MR-Egger (SIMEX) | 0.023 | 0.0110 | $4.5 \times 10^{-2}$ |
| Collider-Corrected LAD | 0.021 | 0.0056 | $1.5 \times 10^{-4}$ |
| Collider-Corrected LAD (SIMEX) | 0.024 | 0.0042 | $1.4 \times 10^{-8}$ |
| Collider-Corrected MR-RAPs | 0.023 | 0.0043 | $3.6 \times 10^{-8}$ |

Across all methods, we see a consistent picture: a unit increase in the log-odds of insomnia leads to an increase of between 0.17 and 0.24 units of log mmol/mol HbA1c. All estimates are further from the null than the collider biased observational association, 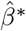. However the results highlight that, without weak-instrument adjustment, all summary data MR-methods are biased in the direction of 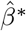.

Table 1 (rows 6:10) and Figure 6 (bottom) show the MR results for analysis using only the 112 SNPs identified in Jansen from 23andMe data, which are immune to the dilution bias caused by winner’s curse. These SNPs have a weaker mean *F* statistic of 6.88 but a higher 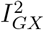 statistic of 52%. All causal estimates are seen to increase when compared to analysis (a). This is because the winner’s curse which is present in (a) leads to an over-estimation of the SNP-exposure association (which forms the denominator of the standard ratio estimate for *β*) and thus an underestimation of the causal effect. Again, across all methods, we see consistent evidence that the insomnia causally increases HbA1c.

In total there were 14 outlier SNPs (13 SNPs in analysis (a) and (6) in analysis (b), respectively), which were investigated using the GWAS Catalog (https://www.ebi.ac.uk/gwas/), a full list of which can be found in *Supplementary Information*. Most of these SNPs are only associated with insomnia except rs10758593 (type 1 and type 2 diabetes), rs12917449 (type 2 diabetes), rs1861412 (BMI) and rs429358 (70+ traits). This provides some biological evidence for the existence of pleiotropy, which further underlines the utility of using robust methods that account for its presence.

## Discussion

In this paper we clarify how the principle of Collider-Correction offers a vehicle for applying any two-sample summary data MR method to one sample data, making it easy to account for both pleiotropy and weak instrument bias. Our method is closely related to the approach of Dudbridge et al [30] for genetic studies of disease progression, and primarily serves to emphasise that this procedure is in fact applicable to any MR analysis. We used our new method to provide important insights into the role of insomnia on glycated haemoglobin and, by extension, on incident diabetes.

A nice feature of our approach is that the Collider-Correction term *β* − *β*^*^ will be large (and therefore the Collider-Corrected estimate will be clearly distinct from the observational association) precisely when there is strong confounding. Conversely, when there is weak confounding, or the confounding has been sufficiently adjusted for, *β* − *β*^*^ will be zero and Collider-Correction estimate will equal the observational association. In this case, the observational association then becomes a consistent and likely very efficient estimate of the true causal effect. Collider-Correction therefore naturally promotes the triangulation and synthesis of observational and MR estimates, which can estimate the true causal effect with distinct but complementary assumptions.

We showcased the Collider-Correction approach using four univariate MR approaches that estimate a single causal effect parameter. At the cutting-edge of MR methods research, new approaches are attempting to: estimate causal effects identified by different clusters of SNPs [49, 50, 31]; simultaneously estimate causal effects via multiple exposures [51, 52], or quantify non-linear effects of an exposure [53]. The Collider-Correction algorithm can in principle be adapted to fit all of these multi-parameter approaches and this is an important topic of future research.

The insomnia data was affected by a small amount of winner’s curse, which we removed by design in a sensitivity analysis by restricting our SNP set to those obtained from a purely independent data source. More sophisticated approaches to adjusting for winner’s curse are possible by incorporating the original Discovery data. For example, Bowden and Dudbridge [4] describe the most statistically efficient way to combine SNP discovery and validation data from two non-overlapping GWAS studies and remove winner’s curse. As further work, we plan to extend this approach and combine it with Collider-Correction.

## Supporting information

R code for simulation study and outlier analysis spreadsheet

## Data Availability

This paper analyses data from the publicly accessible UK Biobank study and published summary statistics. Information on UK Biobank is available at https://www.ukbiobank.ac.uk/principles-of-access/

## Acknowledgement

Ciarrah Barry is supported by a Wellcome Trust studentship (218495/Z/19/Z). James Liu is funded by a Diabetes UK project grant (17/0005700). Deborah A Lawlor, James Liu and Rebecca Richmond all work in a Unit that receives support from the University of Bristol and UK Medical Research Council (MCUU00011/6). Deborah A Lawlor is a National Institute of Research Senior Investigator (NF-0616-10102). Frank Dudbridge is supported by the MRC (MR/S037055/1). Jack Bowden is funded by an Expanding Excellence in England (E3) research award. None of the funders influenced the research presented here and the views expressed in this paper are those of the authors and not necessarily any funders acknowledged here.

## Author contributions

Barry and Bowden developed the Collider-Correction algorithm and its rationale was further refined by Dudbridge. Liu, Richmond and Bowden applied the method to UK Biobank and 23andMe data to produce the results detailed in the paper. Bowden, Barry and Dudbridge produced an initial draft of the paper which was further refined by Lawlor, Rutter, Liu and Richmond. All authors read and approved the final version of the manuscript.

## Supplementary information

### A Derivation of Equation (6)

The asymptotic least squares estimates of the effects of *X* and *G* on *Y*, without conditioning on *U*, are

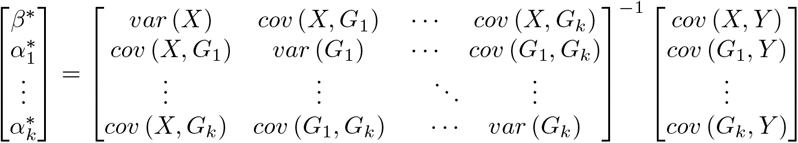

Assuming no LD between SNPs, so *cov* (*G*_*i*_, *G*_*j*_) = 0 where *i* ≠ *j*, the variance-covariance matrix has block form with a diagonal matrix in the lower right quadrant. Block-wise inversion gives

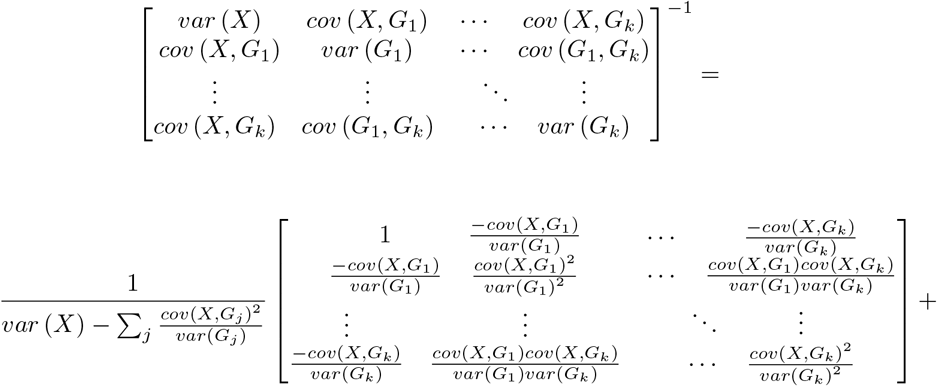

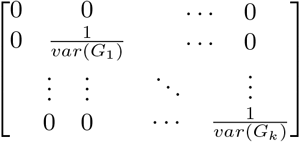

Then

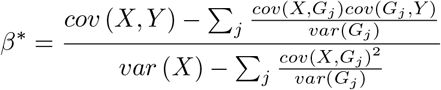

And

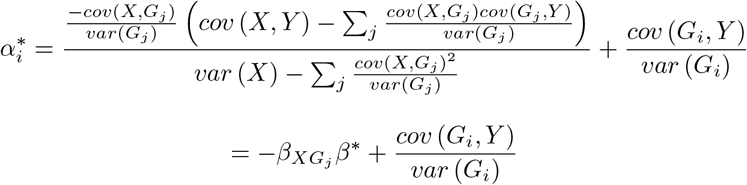

From equation 2, 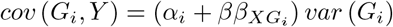. Therefore

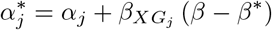

The causal effect *β* is therefore the observational effect *β*^*^, plus the slope of the regression of 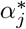 on 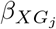.

### B Standard deviation plots for Section 3 simulation study

### C Mean-Squared Error plots for Section 3 simulation study

### D Outlier SNPs sets for the Insomnia analysis of Section 4

SNP set detected as outliers using a Bonferroni corrected exact *Q* statistic (23andMe data only)

|  |  |
| --- | --- |
| 1 | rs10758593 |
| 2 | rs1264419 |
| 3 | rs214934 |
| 4 | rs2792990 |
| 5 | rs4788203 |
| 6 | rs647905 |

SNP set detected as outliers using a Bonferroni corrected exact *Q* statistic (23andMe + UK Biobank data)

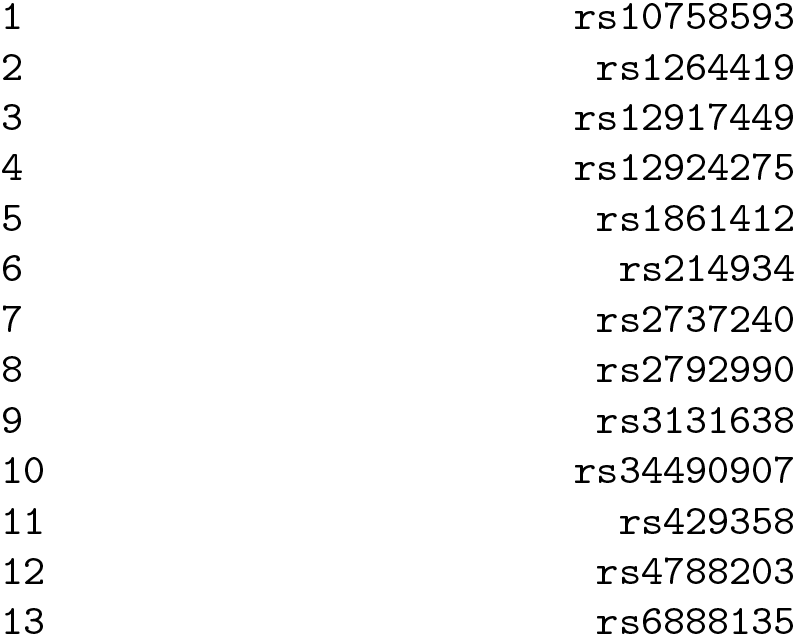

