## Supplementary material for "Exploiting collider bias to apply two-sample summary data Mendelian randomization methods to one-sample individual level data": R code for simulation study and outlier analysis spreadsheet: ColliderCorrectionPaperCode.R

```
# IVW simulation

library(simex)
#Simulation


DataFile     = "IVWData.RData"
MAINUNCOR    = matrix(nrow=10,ncol=12)
MAINUNCOR.SD = matrix(nrow=10,ncol=12)

for(j in 1:10){
  
  N       = 5000*j                   # variable sample size
  beta_UX = 3
  beta_UY = 2
  beta    = 0.5                      # true value of beta we wish to recover
  n       = N                        # sample size
  size    = 50                       # number of genetic variants  
  prob    = 0.3                      # allele frequency
  Sims    = 1000                      # number of simulations per sample size
  sdU     = 5
  
  Results.2S = matrix(nrow=Sims,ncol=12)
  V1 = V2 = V3 = NULL
  
  for(i in 1:Sims){
    
    
    BXG             = matrix(runif(size,0.1,0.5),nrow=size)            # coefficients for genetic variants
    G1.2            = matrix(rbinom(n*size,2,prob),n,size)             # number of alleles
    U.2             = rnorm(n,0,sdU)                                   # independent confounding 
    Ex.2            = rnorm(n,0,4)                                     # independent residual error
    X.2             = G1.2%*%BXG  + beta_UX*U.2 + Ex.2                 # X~G, generate the data
    Ey.2            = rnorm(n,0,2)                                     # independent residual error
    beta_GY1        = matrix(rnorm(size,0,0.2),nrow=size)              # balanced pleiotropy
    Y               = G1.2%*%beta_GY1  + beta*X.2 + beta_UY*U.2 +Ey.2  # Y~G+X,generate the data
    OBS             = summary(lm(Y~X.2))$coef[2]
    
    #  True confounder effect
    
    Coll           = beta_UX*beta_UY*sdU^2
    
    
    XGdata2        = data.frame(X.2,G1.2)             
    FIT2           = summary(lm(XGdata2))             # estimated linear association
    BetaXG         = FIT2$coef[-1,1]                  # estimated association for genetic variants, X~G
    seBetaXG       = FIT2$coef[-1,2]                  # standard error for estimated association for genetic variants
    V1             = BetaXG-BXG        
    Fbar           = mean((BetaXG^2)/(seBetaXG^2))             # f-statistic
    
    
    # Y~G  (for standard IVW)
    
    YGdata         = data.frame(Y,G1.2) 
    FIT3           = summary(lm(YGdata))               # estimated linear association 
    BetaYG         = FIT3$coef[-1,1]                   # estimated association between outcome and genetic variants
    seBetaYG       = FIT3$coef[-1,2]                   # standard error for estimated association for genetic variants
    V2             = BetaYG-beta_GY1-beta*BXG
    
    # Y~G+X (two-step IVW)
    
    YXGdata          = data.frame(Y,X.2,G1.2)
    FIT4             = summary(lm(YXGdata))                  # estimated associations
    alphahatstar     = FIT4$coef[-c(1,2),1]                  # obtain the estimates for the genetic variant coefficient,
    # we have the analytic expression for these estimates
    se.alphahatstar  = FIT4$coef[-c(1,2),2]
    betastar         = FIT4$coef[2,1]                        # obtain the estimate for the exposure of interest, we have 
    # the analytic expression for the estimate
        
    # Standard 1 sample IVW
    
    MRE1 = summary(lm(BetaYG~-1+BetaXG),weights=1/seBetaYG^2)$coef[1,1]
    
    # Simex 1 sample IVW
    
    Fit       = lm(BetaYG~-1+BetaXG,weights=1/seBetaYG^2,x=TRUE,y=TRUE)
    mod.sim1  = simex(Fit,B=250,measurement.error=seBetaXG,SIMEXvariable="BetaXG",fitting.method="quad",asymptotic="FALSE")
    MRE2      = summary(mod.sim1)$coef$jackknife[1]                  # obtain the SIMEX corrected estimate
    
    # Standard one sample two step IVW
    
    A             = alphahatstar                                  # multiply up by the denominator of the RHS of the analytic expression
    betahat       = summary(lm(A~ -1 + BetaXG))$coef[1,1]           # regress the modified alphahatstar on BetaXG
    MRE3          = betastar + betahat                             # equation rearrangement to recover the adjusted estimate for beta
    
    # SIMEX one sample two step IVW 
    
    Fit       = lm(A~ -1+ BetaXG,x=TRUE,y=TRUE)                    # regress the modified alphahatstar on betaXG
    mod.sim2  = simex(Fit,B=250,measurement.error=seBetaXG,SIMEXvariable="BetaXG",fitting.method="quad",asymptotic="FALSE")
    bSIMEX    = summary(mod.sim2)$coef$jackknife[1]                # obtain the SIMEX corrected estimate
    MRE4      = betastar + bSIMEX                 # equation rearrangement to recover the adjusted estimate for beta
    
    
    # TSLS
    
    Xhat           = lm(XGdata2)$fitted
    TSLS           = summary(lm(Y~Xhat))$coef[2]
    
    # two sample approach (split data in two)
    
    n1             = n/2
    
    X.21           = X.2[1:n1]
    G1.21          = G1.2[1:n1,]
    n2             = n1+1  
    G1.22          = G1.2[n2:n,]  
    Y.2            = Y[n2:n]
 
    
    XGdata2        = data.frame(X.21,G1.21)             
    FIT2           = summary(lm(XGdata2))             # estimated linear association
    BetaXG         = FIT2$coef[-1,1]                  # estimated association for genetic variants, X~G
    seBetaXG       = FIT2$coef[-1,2]                  # standard error for estimated association for genetic variants    
    YGdata         = data.frame(Y.2,G1.22) 
    FIT3           = summary(lm(YGdata))               # estimated linear association 
    BetaYG         = FIT3$coef[-1,1]                   # estimated association between outcome and genetic variants
    seBetaYG       = FIT3$coef[-1,2]                   # standard error for estimated association for genetic variants
    
    TwoSIVW = summary(lm(BetaYG~-1+BetaXG,weights=1/seBetaYG^2))$coef[1,1]
    
    Fit       = lm(BetaYG~-1+BetaXG,weights=1/seBetaYG^2,x=TRUE,y=TRUE)                    # regress the modified alphahatstar on betaXG
    mod.sim2  = simex(Fit,B=250,measurement.error=seBetaXG,SIMEXvariable="BetaXG",fitting.method="quad",asymptotic="FALSE")
    TwoSIVW_S = summary(mod.sim2)$coef$jackknife[1]                # obtain the SIMEX corrected estimate
          
    
    
    Results.2S[i,1]  = MRE1          # standard 1 sample IVW 
    Results.2S[i,2]  = MRE2          # SIMEX    1 sample IVW 
    Results.2S[i,3]  = MRE3          # Standard 1 sample two step IVW 
    Results.2S[i,4]  = MRE4          # SIMEX    1 sample two step IVW 
    Results.2S[i,5]  = betahat       # Standard one sample two step IVW regression
    Results.2S[i,6]  = Fbar          # F-statistic
    Results.2S[i,7]  = OBS           # Standard Observational Estimate
    Results.2S[i,8]  = TSLS          # TSLS
    Results.2S[i,9]  = cor(V1,V2)    # Correlation between G-X,G-Y assoc. 
    Results.2S[i,10] = cor(V1,V3)    # Correlation between G-X & G-Y|X assoc.
    Results.2S[i,11] = TwoSIVW       # 
    Results.2S[i,12] = TwoSIVW_S    # 
    
    
    print(i)
    print(j)
    
  }
  
  MAINUNCOR[j,]    = apply(Results.2S,2,mean,na.rm=TRUE)
  MAINUNCOR.SD[j,] = apply(Results.2S,2,sd,na.rm=TRUE)
  
  
  
}


SS                = 5000*seq(1,10)


save(MAINUNCOR,MAINUNCOR.SD,file=DataFile)


library(simex)
#setwd()


# MR Egger simulation


DataFile = "MREggerData.RData"


Isq = function(y,s)
{
  k          = length(y)
  w          = 1/s^2; sum.w  = sum(w)
  mu.hat     = sum(y*w)/sum.w  
  Q          = sum(w*(y-mu.hat)^2)
  Isq        = (Q - (k-1))/Q
  Isq        = max(0,Isq)
  return(Isq)
}

#Simulation

MAIN.MRE    = matrix(nrow=10,ncol=7)
MAIN.MRE.SD = matrix(nrow=10,ncol=7)

for(j in 1:10){
  
  N                = 5000*j  
  beta_UX = 3
  beta_UY = 2
  beta    = 0.5                      # the true value of beta we wish to recover
  n       = N                        # sample size
  size    = 50                       # number of genetic variants  
  prob    = 0.3                      # allele frequency
  Sims    = 1000                      # number of simulations per sample size
  sdU     = 5
  
  Results.2S     = matrix(nrow=Sims,ncol=7)
  V1             = V2 = V3 = NULL
  
  for(i in 1:Sims){
    

    
    BXG             = matrix(runif(size,0.1,0.5),nrow=size)            # coefficients for genetic variants
    G1.2            = matrix(rbinom(n*size,2,prob),n,size)             # number of alleles
    U.2             = rnorm(n,0,sdU)                                   # independent confounding
    Ex.2            = rnorm(n,0,4)                                     # independent residual error
    X.2             = G1.2%*%BXG  + beta_UX*U.2 + Ex.2                 # X~G, generate the data
    Ey.2            = rnorm(n,0,2)                                     # independent residual error
    beta_GY1        = matrix(rnorm(size,0.1,0.1),nrow=size)              # directional pleiotropy
    Y               = G1.2%*%beta_GY1  + beta*X.2 + beta_UY*U.2 +Ey.2  # Y~G+X,generate the data
    OBS             = summary(lm(Y~X.2))$coef[2]
    
    #  True confounder effect
    
    Coll = beta_UX*beta_UY*sdU^2
    
    # X~G
    
    XGdata2        = data.frame(X.2,G1.2)
    FIT2           = summary(lm(XGdata2))                              # estimated linear association
    BetaXG         = FIT2$coef[-1,1]                                   # estimated association for genetic variants, X~G
    seBetaXG       = FIT2$coef[-1,2]                                   # standard error for estimated association for genetic variants
   
    # Y~G  (for standard MR-Egger)
    
    YGdata         = data.frame(Y,G1.2)
    FIT3           = summary(lm(YGdata))                               # estimated linear association                         
    BetaYG         = FIT3$coef[-1,1]                                   # estimated association between outcome and genetic variants
    seBetaYG       = FIT3$coef[-1,2]                                   # standard error for estimated association for genetic variants
    
    # Y~G+X (two-step MR-Egger)
    
    YXGdata          = data.frame(Y,X.2,G1.2)
    FIT4             = summary(lm(YXGdata))                           # estimated associations
    alphahatstar     = FIT4$coef[-c(1,2),1]                           # obtain the estimates for the genetic variant coefficient,
                                                                      # we have the analytic expression for these estimates
    se.alphahatstar  = FIT4$coef[-c(1,2),2]
    betastar         = FIT4$coef[2,1]                                 # obtain the estimate for the exposure of interest, we have 
                                                                      # the analytic expression for the estimate
    
    A                = alphahatstar
    betahat          = summary(lm(A~ +BetaXG))$coef[2,1]
    
    
    # Standard 1 sample MR-Egger
    
    MRE1 = summary(lm(BetaYG~BetaXG),weights=1/seBetaYG^2)$coef[2,1]
  
    # Simex 1 sample MR egger
    
    Fit       = lm(BetaYG~BetaXG,weights=1/seBetaYG^2,x=TRUE,y=TRUE)
    mod.sim1  = simex(Fit,B=250,measurement.error=seBetaXG,SIMEXvariable="BetaXG",fitting.method="quad",asymptotic="FALSE")
    MRE2      = summary(mod.sim1)$coef$jackknife[2]                    # obtain the SIMEX corrected estimate
    
    # Standard one sample two step MR-Egger 
    
    A             = alphahatstar                                   # multiply up by the denominator of the RHS of the analytic expression
    betahat       = summary(lm(A~ BetaXG))$coef[2,1]           # regress the modified alphahatstar on BetaXG
    MRE3          = betastar + betahat    # equation rearrangement to recover the adjusted estimate for beta
    I2betahat     = Isq(BetaXG,seBetaXG)                               # I-squared statistic
    
    # SIMEX one sample two step MR-Egger 
    
    Fit       = lm(A~ BetaXG,x=TRUE,y=TRUE)                    # regress the modified alphahatstar on betaXG
    mod.sim2  = simex(Fit,B=250,measurement.error=seBetaXG,SIMEXvariable="BetaXG",fitting.method="quad",asymptotic="FALSE")
    bSIMEX    = summary(mod.sim2)$coef$jackknife[2]                    # obtain the SIMEX corrected estimate
    MRE4      = betastar + bSIMEX        # equation rearrangement to recover the adjusted estimate for beta
    
    
    Results.2S[i,1]  = MRE1          # Standard 1 sample MRE 
    Results.2S[i,2]  = MRE2          # SIMEX    1 sample MRE 
    Results.2S[i,3]  = MRE3          # Standard 1 sample two step MRE 
    Results.2S[i,4]  = MRE4          # SIMEX    1 sample two step MRE 
    Results.2S[i,5]  = betahat       # Standard 1 sample MRE regression
    Results.2S[i,6]  = I2betahat     # I-squared statistic
    Results.2S[i,7]  = OBS           # Standard Observational Estimate
      
         
  }
  
  MAIN.MRE[j,]    = apply(Results.2S,2,mean,na.rm=TRUE)
  
  MAIN.MRE.SD[j,] = apply(Results.2S,2,sd,na.rm=TRUE)
  
  print(j)
 
   
}


save(MAIN.MRE,MAIN.MRE.SD,file=DataFile)


# Weighted median/LAD regression simulation

library(L1pack)
library(simex)

DataFile = "LADRegDataNoInSIDE.RData"

set.seed(1)

#Simulation

MAIN.WghtMed    = matrix(nrow=10,ncol=8)
MAIN.WghtMed.SD = matrix(nrow=10,ncol=8)
BootSE          = matrix(nrow=10,ncol=2)

for(j in 1:10){
  
  
  N       = 5000*j
  beta_UX = 3
  beta_UY = 2
  beta    = 0.5    # true value of beta we wish to recover
  n       = N      # sample size per iteration
  size    = 50     # number of genetic variants
  prob    = 0.3    # allele frequency
  Sims    = 1000    # number of simulations per sample size
  sdU     = 5
  
  
  Results.2S = matrix(nrow=Sims,ncol=8)
  BootSE     = matrix(nrow=Sims,ncol=1)
  V1         = V2 = V3 = NULL
  
  for(i in 1:Sims){

        
    BXG             = matrix(runif(size,0.1,0.5),nrow=size)            # coefficients for genetic variants
    G1.2            = matrix(rbinom(n*size,2,prob),n,size)             # number of alleles
    U.2             = rnorm(n,0,sdU)                                   # independent confounding 
    Ex.2            = rnorm(n,0,4)                                     # independent residual error
    X.2             = G1.2%*%BXG  + beta_UX*U.2 + Ex.2                 # X~G, generate the data
    Ey.2            = rnorm(n,0,2)                                     # independent residual error
    bgy             = rnorm(15,0.2,0.2) + BXG[1:15]
    beta_GY1        = matrix(c(bgy,rep(0,35)),nrow=size)               # unbalanced pleiotropy violating InSIDE!!!
    Y               = G1.2%*%beta_GY1  + beta*X.2 + beta_UY*U.2 +Ey.2  # Y~G+X,generate the data
    OBS             = summary(lm(Y~X.2))$coef[2]


    #  True confounder effect
    
    Coll = beta_UX*beta_UY*sdU^2
    
    # X~G
    
    
    XGdata2        = data.frame(X.2,G1.2)
    FIT2           = summary(lm(XGdata2))               # estimated linear association
    BetaXG         = FIT2$coef[-1,1]                    # estimated association for genetic variants, X~G
    seBetaXG       = FIT2$coef[-1,2]                    # standard error for estimated association for genetic variants
    V1             = BetaXG-BXG                         # difference between true and estimated values for genetic variants
    Fbar           = mean((BetaXG^2)/(seBetaXG^2))      # f-statistic
    
    
    # Y~G 
    
    YGdata         = data.frame(Y,G1.2)
    FIT3           = summary(lm(YGdata))        # estimated linear association
    BetaYG         = FIT3$coef[-1,1]            # estimated association between outcome and genetic variants
    seBetaYG       = FIT3$coef[-1,2]            # standard error for estimated association for genetic variants
    V2             = BetaYG-beta_GY1-beta*BXG   # difference between true and estimated values for genetic variants association
    
    # Y~G+X
    
    YXGdata          = data.frame(Y,X.2,G1.2)
    FIT4             = summary(lm(YXGdata))                                  # estimated associations
    alphahatstar     = FIT4$coef[-c(1,2),1]                                  # obtain the estimates for the genetic variant coefficient,
                                                                             # we have the analytic expression for these estimates
    se.alphahatstar  = FIT4$coef[-c(1,2),2]
    betastar         = FIT4$coef[2,1]                                        # obtain the estimate for the exposure of interest, we have 
                                                                             # the analytic expression for the estimate
       
    
    # Standard 1 sample LAD regression
    
    WMED1   = try(lad(BetaYG~-1+BetaXG,model=TRUE,x=TRUE,y=TRUE)$coef)

    # Simex 1 sample LAD regression
        
    Fit        = try(lad(BetaYG~-1+BetaXG,model=TRUE,x=TRUE,y=TRUE))
    
    WMED2 = NA
    mod.sim1   = try(simex(Fit,B=500,measurement.error=seBetaXG,SIMEXvariable="BetaXG",
                 asymptotic="FALSE",jackknife.estimation = F)$coef)
    
    if(is.numeric(mod.sim1)==TRUE){WMED2      = mod.sim1}                                               # obtain the SIMEX corrected estimate
   
    # Standard one sample two step LAD regression
    
    A             = alphahatstar                                           # multiply up by the denominator of the RHS of the analytic expression
    betahat       = summary(lad(A~ -1 + BetaXG))$coef[1,1]                    # regress the modified alphahatstar on BetaXG
    WMED3         = betastar + betahat        # equation rearrangement to recover the adjusted estimate for beta

    # SIMEX one sample two step weighted-median 
    
    Fit       = try(lad(A~-1+BetaXG,model=TRUE,x=TRUE,y=TRUE))            # regress the modified alphahatstar on betaXG
    
    WMED4 = NA
    mod.sim2   = try(simex(Fit,B=500,measurement.error=seBetaXG,SIMEXvariable="BetaXG",
                           asymptotic="FALSE",jackknife.estimation = F)$coef)
    
    if(is.numeric(mod.sim2)==TRUE){bSIMEX      = mod.sim2
                                   WMED4     = betastar + bSIMEX    }         
    
    
                                                                  # obtain the SIMEX corrected estimate
    
    
    
    # SIMEX one sample two step IVW
    
    Fit       = try(lm(A~-1+BetaXG,model=TRUE,x=TRUE,y=TRUE))            # regress the modified alphahatstar on betaXG
    
    IVW = NA
    mod.sim3   = try(simex(Fit,B=500,measurement.error=seBetaXG,SIMEXvariable="BetaXG",
                           asymptotic="FALSE",jackknife.estimation = F)$coef)
    
    if(is.numeric(mod.sim3)==TRUE){bSIMEX      = mod.sim3
                                   IVW     = betastar + bSIMEX    }         
    
    
    

    
    Results.2S[i,1]  = WMED1          # standard 1 sample weighted-median 
    
    Results.2S[i,2]  = WMED2          # SIMEX    1 sample weighted-median 
    
    Results.2S[i,3]  = WMED3          # Standard 1 sample two step weighted-median 
    
    Results.2S[i,4]  = WMED4          # SIMEX    1 sample two step weighted-median 
    
    Results.2S[i,5]  = betahat        # Standard 1 sample weighted-median
    
    Results.2S[i,6]  = Fbar           # Standard 1 sample two step weighted-median
    
    Results.2S[i,7]  = OBS            # Standard Observational Estimate
    
    Results.2S[i,8]  = IVW            # SIMEX   1 sample two step IVW

    print(i)
    print("new")
    
  }
  
  MAIN.WghtMed[j,]    = apply(Results.2S,2,mean,na.rm=TRUE)
  MAIN.WghtMed.SD[j,] = apply(Results.2S,2,sd,na.rm=TRUE)
  print(j)
  
}

SS                = 5000*seq(1,10)

## graphical results


save(MAIN.WghtMed,MAIN.WghtMed.SD,file=DataFile)


library(simex)

# 1 versus 2 sample IVW simulation 

DataFile     = "IVWData2.RData"
MAINUNCOR2    = matrix(nrow=10,ncol=2)
MAINUNCOR2.SD = matrix(nrow=10,ncol=2)

for(j in 1:10){
  
  N       = 5000*j                   # variable sample size
  beta_UX = 3
  beta_UY = 2
  beta    = 0.5                      # true value of beta we wish to recover
  n       = N                        # sample size
  size    = 50                       # number of genetic variants  
  prob    = 0.3                      # allele frequency
  Sims    = 1000                      # number of simulations per sample size
  sdU     = 5
  
  Results.2S = matrix(nrow=Sims,ncol=2)
  V1 = V2 = V3 = NULL
  
  for(i in 1:Sims){
    
    
    BXG             = matrix(runif(size,0.1,0.5),nrow=size)            # coefficients for genetic variants
    G1.2            = matrix(rbinom(n*size,2,prob),n,size)             # number of alleles
    U.2             = rnorm(n,0,sdU)                                   # independent confounding 
    Ex.2            = rnorm(n,0,4)                                     # independent residual error
    X.2             = G1.2%*%BXG  + beta_UX*U.2 + Ex.2                 # X~G, generate the data
    Ey.2            = rnorm(n,0,2)                                     # independent residual error
    beta_GY1        = matrix(rnorm(size,0,0.2),nrow=size)              # balanced pleiotropy
    Y               = G1.2%*%beta_GY1  + beta*X.2 + beta_UY*U.2 +Ey.2  # Y~G+X,generate the data
    
    
    #  True confounder effect
    
    Coll           = beta_UX*beta_UY*sdU^2
    
    
   
    # two sample approach (split data in two)
    
    n1             = n/2
    
    X.21           = X.2[1:n1]
    G1.21          = G1.2[1:n1,]
    n2             = n1+1  
    G1.22          = G1.2[n2:n,]  
    Y.2            = Y[n2:n]
 
    
    XGdata2        = data.frame(X.21,G1.21)             
    FIT2           = summary(lm(XGdata2))             # estimated linear association
    BetaXG         = FIT2$coef[-1,1]                  # estimated association for genetic variants, X~G
    seBetaXG       = FIT2$coef[-1,2]                  # standard error for estimated association for genetic variants    
    YGdata         = data.frame(Y.2,G1.22) 
    FIT3           = summary(lm(YGdata))               # estimated linear association 
    BetaYG         = FIT3$coef[-1,1]                   # estimated association between outcome and genetic variants
    seBetaYG       = FIT3$coef[-1,2]                   # standard error for estimated association for genetic variants
    
    TwoSIVW = summary(lm(BetaYG~-1+BetaXG,weights=1/seBetaYG^2))$coef[1,1]
    
    Fit       = lm(BetaYG~-1+BetaXG,weights=1/seBetaYG^2,x=TRUE,y=TRUE)                    # regress the modified alphahatstar on betaXG
    mod.sim2  = simex(Fit,B=250,measurement.error=seBetaXG,SIMEXvariable="BetaXG",fitting.method="quad",asymptotic="FALSE")
    TwoSIVW_S = summary(mod.sim2)$coef$jackknife[1]                # obtain the SIMEX corrected estimate
          
    
    Results.2S[i,1] = TwoSIVW       # 
    Results.2S[i,2] = TwoSIVW_S    # 
    
    
    #print(i)
    print(j)
    
  }
  
  MAINUNCOR2[j,]    = apply(Results.2S,2,mean,na.rm=TRUE)
  MAINUNCOR2.SD[j,] = apply(Results.2S,2,sd,na.rm=TRUE)
  
  
  
}


SS                = 5000*seq(1,10)


save(MAINUNCOR2,MAINUNCOR2.SD,file=DataFile)


# MR RAPS simulation

library(simex)
#Simulation

rm(list=ls(all=TRUE))
library(mr.raps)


DataFile     = "MRRAPS.RData"

MAINUNCOR    = matrix(nrow=10,ncol=1)
MAINUNCOR.SD = matrix(nrow=10,ncol=1)
CorObsCC     = matrix(nrow=10,ncol=1)
CorObsCC2     = matrix(nrow=10,ncol=1)


for(j in 1:10){
  
  N       = 5000*j                   # variable sample size
  beta_UX = 3
  beta_UY = 2
  beta    = 0.5                      # true value of beta we wish to recover
  n       = N                        # sample size
  size    = 50                       # number of genetic variants  
  prob    = 0.3                      # allele frequency
  Sims    = 500                      # number of simulations per sample size
  sdU     = 5
  
  Results.2S = matrix(nrow=Sims,ncol=1)
  V1 = V2 = V3 = V4 = bstar =  NULL
  
  for(i in 1:Sims){
    
    
    BXG             = matrix(runif(size,0.1,0.5),nrow=size)            # coefficients for genetic variants
    G1.2            = matrix(rbinom(n*size,2,prob),n,size)             # number of alleles
    U.2             = rnorm(n,0,sdU)                                   # independent confounding 
    Ex.2            = rnorm(n,0,10)# rnorm(n,0,4)                                     # independent residual error
    X.2             = G1.2%*%BXG  + beta_UX*U.2 + Ex.2                 # X~G, generate the data
    Ey.2            = rnorm(n,0,10)#rnorm(n,0,2)                                     # independent residual error
    beta_GY1        = matrix(rnorm(size,0,0.2),nrow=size)              # balanced pleiotropy
    Y               = G1.2%*%beta_GY1  + beta*X.2 + beta_UY*U.2 +Ey.2  # Y~G+X,generate the data
    OBS             = summary(lm(Y~X.2))$coef[2]
    
    #  True confounder effect
    
    Coll           = beta_UX*beta_UY*sdU^2
    
    # X~G
    
    XGdata2        = data.frame(X.2,G1.2)             
    FIT2           = summary(lm(XGdata2))             # estimated linear association
    BetaXG         = FIT2$coef[-1,1]                  # estimated association for genetic variants, X~G
    seBetaXG       = FIT2$coef[-1,2]                  # standard error for estimated association for genetic variants
    V1             = BetaXG-BXG        
    Fbar           = mean((BetaXG^2)/(seBetaXG^2))             # f-statistic
    
    
    # Y~G  (for standard IVW)
    
    YGdata         = data.frame(Y,G1.2) 
    FIT3           = summary(lm(YGdata))               # estimated linear association 
    BetaYG         = FIT3$coef[-1,1]                   # estimated association  between outcome and genetic variants
    seBetaYG       = FIT3$coef[-1,2]                   # standard error for estimated association for genetic variants
    V2             = BetaYG-beta_GY1-beta*BXG
    
    # Y~G+X (two-step IVW)
    
    YXGdata          = data.frame(Y,X.2,G1.2)
    FIT4             = summary(lm(YXGdata))                  # estimated associations
    alphahatstar     = FIT4$coef[-c(1,2),1]                  # obtain the estimates for the genetic variant coefficient,
    se.alphahatstar  = FIT4$coef[-c(1,2),2]
    betastar         = FIT4$coef[2,1]                        # obtain the estimate for the exposure of interest, we have 
    bstar[i]         = betastar    
        
    # MR-RAPS


    braps            =  mr.raps.all(BetaXG,alphahatstar, seBetaXG, se.alphahatstar)
    bRAPs            =  betastar + braps$beta.hat[6]    
    Results.2S[i]    =  bRAPs 

              
    
    print(i)
  
    
  }
  
  print(j)
  MAINUNCOR[j,]    = apply(Results.2S,2,mean,na.rm=TRUE)
  MAINUNCOR.SD[j,] = apply(Results.2S,2,sd,na.rm=TRUE)
  CorObsCC[j,]  = cor(bstar,Results.2S)
  CorObsCC2[j,]  = summary(lm(bstar~Results.2S))$coef[2,4]

    
}

MAINUNCOR.RAPS = MAINUNCOR
MAINUNCOR.RAPSsd = MAINUNCOR.SD

CorObsCC
CorObsCC2

save(MAINUNCOR.RAPS,MAINUNCOR.RAPSsd,CorObsCC,CorObsCC2,file=DataFile)


# Code to show residual error independence property of collider-correction

library(simex)
#Simulation

###################
# IVW simulation  #
###################

V1 = V2 = V3 = NULL

  
  N       = 10000                   # variable sample size
  beta_UX = 3
  beta_UY = 2
  beta    = 0.5                      # true value of beta we wish to recover
  n       = N                        # sample size
  size    = 50                       # number of genetic variants  
  prob    = 0.3                      # allele frequency
  Sims    = 500                      # number of simulations per sample size
  sdU     = 5
  
  Results.2S = matrix(nrow=Sims,ncol=12)
  V1 = V2 = V3 = NULL
  
  for(i in 1:Sims){
    
    
   BXG             = matrix(runif(size,0.1,0.5),nrow=size)            # coefficients for genetic variants
    G1.2            = matrix(rbinom(n*size,2,prob),n,size)             # number of alleles
    U.2             = rnorm(n,0,sdU)                                   # independent confounding 
    Ex.2            = rnorm(n,0,50)                                     # independent residual error
    X.2             = G1.2%*%BXG  + beta_UX*U.2 + Ex.2                 # X~G, generate the data
    Ey.2            = rnorm(n,0,50)                                     # independent residual error
    beta_GY1        = matrix(rnorm(size,0,0.2),nrow=size) +0.9*BXG     # balanced pleiotropy
    Y               = G1.2%*%beta_GY1  + beta*X.2 + beta_UY*U.2 +Ey.2  # Y~G+X,generate the data
    OBS             = summary(lm(Y~X.2))$coef[2]
   


    XGdata2        = data.frame(X.2,G1.2)             
    FIT2           = summary(lm(XGdata2))             # estimated linear association
    BetaXG         = FIT2$coef[-1,1]                  # estimated association for genetic variants, X~G
    seBetaXG       = FIT2$coef[-1,2]   

    YXGdata          = data.frame(Y,X.2,G1.2)
    FIT4             = summary(lm(YXGdata))                  # estimated associations
    alphahatstar     = FIT4$coef[-c(1,2),1]               # estimated linear association 
    betaS            = FIT4$coef[2]               

    V1[i]             = cor(BetaXG-BXG,alphahatstar-beta_GY1-(beta-betaS)*BXG)   


    YGdata         = data.frame(Y,G1.2) 
    FIT3           = summary(lm(YGdata))               # estimated linear association 
    BetaYG         = FIT3$coef[-1,1]    

    V2[i] =        cor(BetaXG-BXG,BetaYG-beta_GY1-beta*BXG) 
    
    V3[i] = cor(beta_GY1,BXG)

 
      print(i)
    
  }

par(mfrow=c(1,2) ) 


plot(BXG,beta_GY1,ylab=expression(alpha[j]),xlab=expression(beta[XGj]),cex.lab=1.7,pch=3,cex=4)
lines(BXG,lm(beta_GY1~BXG)$fitted,lty=2)

plot(V1,pch=19,ylim=range(V1,V2,1),xlab="Simulation",ylab="Residual Correlation",cex.lab=1.3)
mean(V1)
points(V2, col="red",pch=19)
legend("topright",c("Standard IVW","Collider-Correction"),col=c("red","black"),bty="n",cex=1.3,pch=19)

mean(V3)
mean(V2)
```
